# Work Attendance during Acute Respiratory Illness Before and During the COVID-19 Pandemic, United States, 2018–2022

**DOI:** 10.1101/2023.08.03.23293611

**Authors:** Faruque Ahmed, Mary Patricia Nowalk, Richard K. Zimmerman, Todd Bear, Carlos G. Grijalva, H. Keipp Talbot, Ana Florea, Sara Y. Tartof, Manjusha Gaglani, Michael Smith, Huong Q. McLean, Jennifer P. King, Emily T. Martin, Arnold S. Monto, C. Hallie Phillips, Karen J. Wernli, Brendan Flannery, Jessie R. Chung, Amra Uzicanin

**Affiliations:** Centers for Disease Control and Prevention, Atlanta, Georgia, USA; University of Pittsburgh, Pittsburgh, Pennsylvania, USA; Vanderbilt University Medical Center, Nashville, Tennessee, USA; Kaiser Permanente Southern California, Pasadena, California, USA; Baylor Scott and White Health, Temple Texas, USA; Texas A&M University College of Medicine, Temple, Texas, USA; Marshfield Clinic Research Institute, Marshfield, Wisconsin, USA; University of Michigan, Ann Arbor, Michigan, USA; Kaiser Permanente Washington Health Research Institute, Seattle, Washington, USA

**Keywords:** Acute respiratory illness, COVID-19, influenza, pandemic, presenteeism, telework, transmission, workplace

## Abstract

Severe Acute Respiratory Syndrome Coronavirus 2 (SARS-CoV-2) and influenza viruses can be transmitted by infected persons who are pre-symptomatic or symptomatic. To assess impact of the COVID-19 pandemic on work attendance during illness, we analyzed prospectively collected data from persons with acute respiratory illness (ARI) enrolled in a multi-state study during 2018–2022. Persons with prior experience working from home were significantly less likely than those without this experience to work onsite on the day before illness and during the first 3 days of illness; the effect was more pronounced for the COVID-19 pandemic period than the pre-pandemic influenza seasons. Persons with influenza or COVID-19 were significantly less likely to work onsite than persons with other ARIs. Among persons for whom positive COVID-19 test results were available by the second or third day of illness, few worked onsite. Work-from-home policies may reduce the likelihood of workplace exposures to respiratory viruses.

**Article’s summary line:** Work-from-home policies may reduce the likelihood of workplace exposures to SARS-CoV-2 and influenza viruses.

## Introduction

Coronavirus Disease 2019 (COVID-19) cases in the U.S., which were first reported on January 22, 2020, began to accelerate in March 2020 (1). The pandemic resulted in a substantial number of employed persons being laid-off or furloughed, especially during the spring of 2020, as well as increased prevalence of working from home (2-4). Employers were advised to actively encourage employees with symptoms of acute respiratory illness (ARI) to stay home (5). Severe Acute Respiratory Syndrome Coronavirus 2 (SARS-CoV-2) and influenza viruses can be transmitted by infected persons who are asymptomatic, pre-symptomatic, or symptomatic (6, 7). Staying home during illness can reduce workplace virus transmission by reducing contacts between infectious and healthy persons (8). It is considered an everyday preventive action that should be implemented year-round and is especially important during annual seasonal influenza seasons and during pandemics (9).

Based on data collected during the early COVID-19 pandemic (March 26, 2020– November 5, 2020), we reported that employed adults with prior hybrid work experience were less likely to work onsite while sick than those without prior hybrid work experience (10). However, whether persons worked onsite early during their illness when infectiousness is higher remains unclear (7, 11, 12). Thus, the goal of our present analysis was to assess the effect of the type of work experience before illness onset on work attendance patterns during the first 3 days of illness among persons with ARI before and during the COVID-19 pandemic.

## Methods

### Study Population

Adults ages 19–64 years were enrolled in the U.S. Influenza Vaccine (Flu VE) Effectiveness Network. From November 12, 2018–March 18, 2020, persons seeking care for an ARI with cough within 7 days of illness onset were enrolled after local influenza circulation was identified from outpatient facilities affiliated with five sites in Michigan (Ann Arbor and Detroit); Pennsylvania (Pittsburgh); Texas (Temple and surrounding areas in Central Texas); Washington (Puget Sound region); and Wisconsin (Marshfield, Wausau, and Weston). From October 14, 2020 through June 30, 2022, the case definition was broadened to include those presenting to outpatient (or telehealth) facilities within 10 days of illness onset with cough, fever, loss of taste or smell, or persons seeking clinical COVID-19 testing. Two additional sites (Southern California region and Nashville, Tennessee) participated from October 2021 through June 2022. November 2018–March 2020 represents pre-pandemic influenza seasons, and October 2020– June 2022 represents a COVID-19 pandemic period. Data collected by the five sites during the early part of the COVID-19 pandemic have been presented in a separate manuscript (10). The study methods have been published previously (13-15). The institutional review boards at the Centers for Disease Control and Prevention and all participating sites approved the study. The enrollees provided informed consent.

### Data Collection

Data collected at enrollment throughout the entire study period November 2018–June 2022 included date of illness onset, age, sex, race/ethnicity, education, self-rated general health status, cigarette smoking, and number of children <12 years of age living in household. Respiratory specimens were collected from all participants at enrollment and tested for influenza viruses using real-time reverse transcription PCR; specimens were also tested for SARS-CoV-2 using reverse transcription PCR during the COVID-19 period (2020–2022). Persons enrolled on or after January 15, 2022 were asked if they had taken at-home rapid COVID-19 tests during their illness and whether the test was positive.

All participants were asked to complete a follow-up survey 1–2 weeks after enrollment either online or over the phone. Throughout the 4-year study period, participants were asked at follow-up whether they fully or mostly recovered from their illness, as well as their employment status, type of employment (hourly; salaried; other), hours expected to work and hours usually worked from home in a typical week, and whether their employer discouraged workers with flu-like symptoms from coming to work (Appendix Table 1). They were also asked about if and where they worked during each of the first 3 days of illness. Work status during the day before illness were asked by the Pennsylvania site for November 2018–May 2019, and by all participating sites for the subsequent study years (Appendix Table 2). Two sites, in Washington and Wisconsin, did not collect data for the period November 2018–September 2021 on work status during illness from participants who typically worked solely from home. For the pre-pandemic influenza seasons, participants were asked at follow-up whether they worked in a healthcare setting with direct patient contact; this question was asked at enrollment for the COVID-19 pandemic period.

### Definitions

Responses to the questions on the number of hours expected to work and worked from home in a typical week were used to categorize the work experience before illness onset (Appendix Figure 1). Employed persons who reported that they usually worked zero hours from home were categorized as having only in-person experience. Persons who stated that the hours usually worked from home were less than the total hours expected to work were categorized as having hybrid experience. The remaining persons were categorized as having only work from home (WFH) experience.

Daily work attendance was categorized as scheduled to work, worked, and not worked. Persons who were scheduled to work for any number of hours in a day were categorized as being scheduled to work. Among persons scheduled to work, those who worked for any number of hours in a day were categorized as having worked and the remaining persons were categorized as having not worked (Appendix Figure 2). Among persons who worked, those who reported work location as “at work” or “both at work and remotely” were categorized as having worked onsite.

A positive result from a PCR test for influenza or SARS-CoV-2 virus was classified as laboratory-confirmed influenza or COVID-19, respectively. Persons with negative PCR test results for influenza or SARS-CoV-2 were categorized as having other ARI.

### Assembly of participants

Among 21,133 unique participants, 61% (12,941/21,133) completed the follow-up survey within 28 days of illness onset (Appendix Figure 3). The follow-up survey completion rates were 39% (Texas), 43% (Michigan), 60% (Washington), 75% (California), 75% (Pennsylvania), 79% (Wisconsin), and 89% (Tennessee). Among those who completed the follow-up survey, 69% (8,936/12,941) worked for an employer for ≥20 hours a week prior to their illness. Of these, after excluding persons who had missing information on hours usually worked from home before illness or persons who had indeterminate or missing laboratory results, 91% (8,132/8,936) were included in the analysis.

### Statistical Analysis

Chi-square test was used to assess differences between frequencies, and Wilcoxon’s rank-sum test was used to compare differences in spread and medians (16). Adjusted odds ratios (aOR) for each day were computed by fitting multi-level logistic regression models to account for the clustering of participants within study sites using PROC GLIMMIX in SAS version 9.4 (SAS Institute, https://www.sas.com). Two sets of regressions were run for employed persons who were scheduled to work. For the first set of regressions, the dependent variable was worked at any location. For the second set of regressions, which examined the location of work to assess the potential to infect coworkers, the dependent variable was worked onsite. Because persons with only WFH experience before illness onset were unlikely to work onsite during illness, they were excluded from the analyses pertaining to location of work.

Independent variables for retention in the models were determined using a backward selection process using change in –2 log likelihood to assess model fit. Age, sex, education, and number of children in the household were ultimately dropped from the models. The variables that were included in the models are shown in the relevant tables. Interactions were assessed.

## Results

Among the 8,132 participants eligible for analysis, there were 1,245 persons with influenza and 2,362 persons with other ARI during the pre-pandemic influenza seasons (Appendix Figure 4). There were 114 persons with influenza, 1,888 with COVID-19, and 2,523 with other ARI during the COVID-19 pandemic period.

Among all participants, 14.0% (1,139) had only WFH experience before illness onset, 18.5% (1,503) had hybrid experience, and 67.5% (5,490) had only in-person experience (Appendix Table 3). Among the 1,139 persons with WFH experience, 88.9% were in the COVID-19 pandemic period and 11.1% were in the pre-pandemic influenza seasons. Among persons with hybrid experience, the median hours usually worked from home in a typical week before illness onset was significantly higher in the pandemic period than in the pre-pandemic influenza seasons (16 hours vs. 8 hours, p<0.001). The proportion of hourly workers was significantly lower among persons with WFH (29.9%) or hybrid (21.8%) experience than among persons with in-person only experience (66.6%) (p<0.001). The proportion of participants who were healthcare personnel varied by work experience (WFH: 7.1%, hybrid: 15.5%, in-person: 25.4%, p<0.001). The proportion with at least a bachelor’s degree was significantly higher among persons with WFH (71.3%) or hybrid (79.5%) experience than those with in-person only experience (43.5%) (p<0.001). The median interval from illness onset to enrollment was 4 days for the WFH experience group and 3 days for the hybrid and in-person groups (p<0.001).

About three fourths of the participants were scheduled to work on each day of illness (Appendix Table 3). Among persons who worked, the median hours worked at any location was 8 hours (interquartile range, IQR: 8, 8) for the day before illness and 8 hours (IQR: 6, 8) for each day of illness (Table 1 footnote). Persons with WFH or hybrid experience were significantly more likely to work at any location on the second and third days of illness compared to those with in-person only experience (Table 1). For example, on the third day of illness during the COVID-19-dominated period, the percentage who worked was 72.4% for persons with WFH experience, 65.2% for persons with hybrid experience, and 37.4% for those with in-person only experience (p<0.001).

**Table 1.**
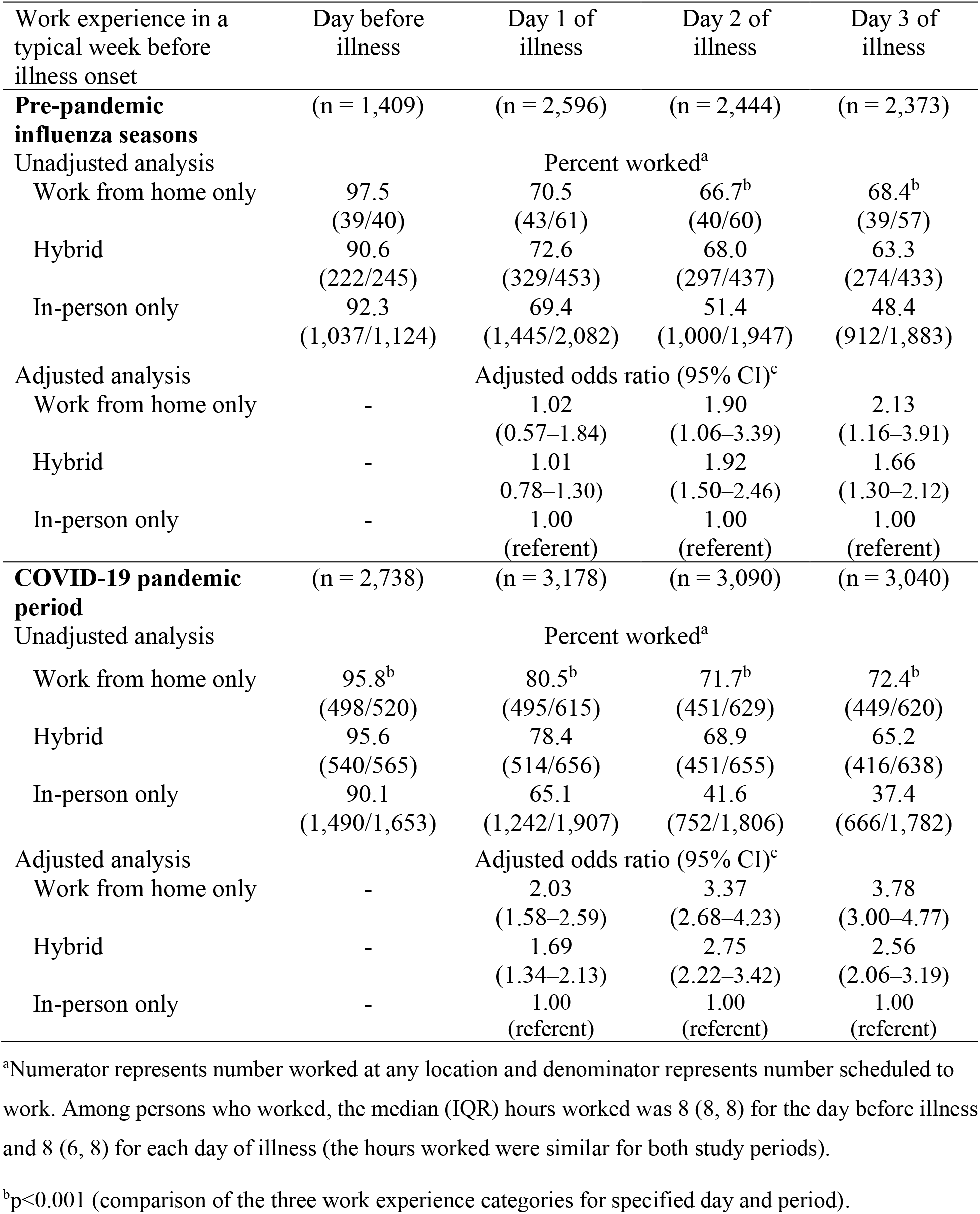
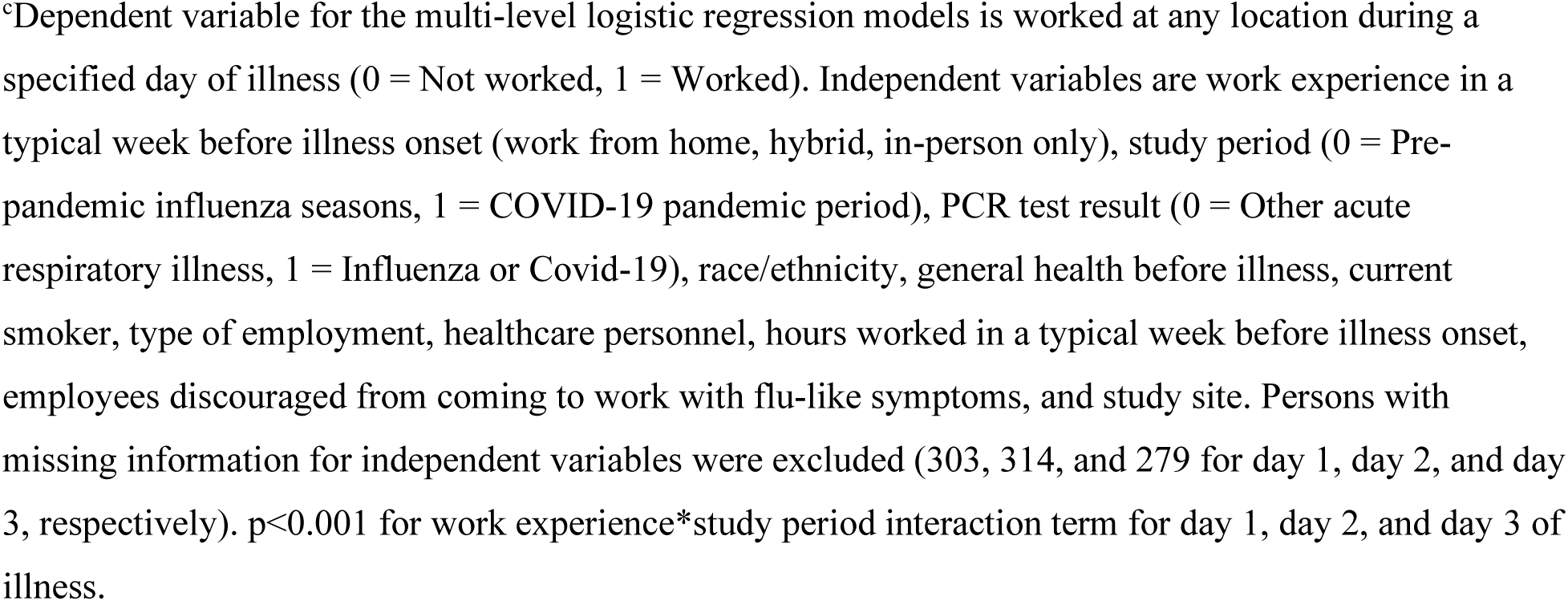
Likelihood of working at any location among adults with COVID-19, influenza, or other acute respiratory illness who were scheduled to work, by prior experience working from home, United States, 2018–2022.

| Work experience in a typical week before illness onset | Day before illness | Day 1 of illness | Day 2 of illness | Day 3 of illness |
| --- | --- | --- | --- | --- |
| **Pre-pandemic influenza seasons** | (n = 1,409) | (n = 2,596) | (n = 2,444) | (n = 2,373) |
| Unadjusted analysis | Percent worked<sup>a</sup> |  |  |  |
| Work from home only | 97.5<br>(39/40) | 70.5<br>(43/61) | 66.7<sup>b</sup><br>(40/60) | 68.4<sup>b</sup><br>(39/57) |
| Hybrid | 90.6<br>(222/245) | 72.6<br>(329/453) | 68.0<br>(297/437) | 63.3<br>(274/433) |
| In-person only | 92.3<br>(1,037/1,124) | 69.4<br>(1,445/2,082) | 51.4<br>(1,000/1,947) | 48.4<br>(912/1,883) |
| Adjusted analysis | Adjusted odds ratio (95% CI)<sup>c</sup> |  |  |  |
| Work from home only | - | 1.02<br>(0.57–1.84) | 1.90<br>(1.06–3.39) | 2.13<br>(1.16–3.91) |
| Hybrid | - | 1.01<br>(0.78–1.30) | 1.92<br>(1.50–2.46) | 1.66<br>(1.30–2.12) |
| In-person only | - | 1.00<br>(referent) | 1.00<br>(referent) | 1.00<br>(referent) |
| **COVID-19 pandemic period** | (n = 2,738) | (n = 3,178) | (n = 3,090) | (n = 3,040) |
| Unadjusted analysis | Percent worked<sup>a</sup> |  |  |  |
| Work from home only | 95.8<sup>b</sup><br>(498/520) | 80.5<sup>b</sup><br>(495/615) | 71.7<sup>b</sup><br>(451/629) | 72.4<sup>b</sup><br>(449/620) |
| Hybrid | 95.6<br>(540/565) | 78.4<br>(514/656) | 68.9<br>(451/655) | 65.2<br>(416/638) |
| In-person only | 90.1<br>(1,490/1,653) | 65.1<br>(1,242/1,907) | 41.6<br>(752/1,806) | 37.4<br>(666/1,782) |
| Adjusted analysis | Adjusted odds ratio (95% CI)<sup>c</sup> |  |  |  |
| Work from home only | - | 2.03<br>(1.58–2.59) | 3.37<br>(2.68–4.23) | 3.78<br>(3.00–4.77) |
| Hybrid | - | 1.69<br>(1.34–2.13) | 2.75<br>(2.22–3.42) | 2.56<br>(2.06–3.19) |
| In-person only | - | 1.00<br>(referent) | 1.00<br>(referent) | 1.00<br>(referent) |

<sup>a</sup>Numerator represents number worked at any location and denominator represents number scheduled to work. Among persons who worked, the median (IQR) hours worked was 8 (8, 8) for the day before illness and 8 (6, 8) for each day of illness (the hours worked were similar for both study periods).
<sup>b</sup>p<0.001 (comparison of the three work experience categories for specified day and period).
<sup>c</sup>Dependent variable for the multi-level logistic regression models is worked at any location during a specified day of illness (0 = Not worked, 1 = Worked). Independent variables are work experience in a typical week before illness onset (work from home, hybrid, in-person only), study period (0 = Pre-pandemic influenza seasons, 1 = COVID-19 pandemic period), PCR test result (0 = Other acute respiratory illness, 1 = Influenza or Covid-19), race/ethnicity, general health before illness, current smoker, type of employment, healthcare personnel, hours worked in a typical week before illness onset, employees discouraged from coming to work with flu-like symptoms, and study site. Persons with missing information for independent variables were excluded (303, 314, and 279 for day 1, day 2, and day 3, respectively). $p < 0.001$ for work experience\*study period interaction term for day 1, day 2, and day 3 of illness.

Analysis of the location of work showed that participants were significantly more likely to work remotely on the day before illness and during the first 3 days of illness during the COVID-19 pandemic period compared with the pre-pandemic influenza seasons (Table 2). For example, on the third day of illness, 18.5% of persons worked remotely during the pandemic period compared with 8.8% during the pre-pandemic influenza seasons.

**Table 2.** Reported work location among adults with influenza, COVID-19, or other acute respiratory illness who were scheduled to work, United States, 2018-2022^a^.

| Location of work | Day before<br>illness | Day 1 of<br>illness | Day 2 of<br>illness | Day 3 of<br>illness |
| --- | --- | --- | --- | --- |
|  | No. (column percent) |  |  |  |
| **Pre-pandemic influenza seasons** | (n = 1,358) | (n = 2,515) | (n = 2,372) | (n = 2,304) |
| At work | 1,161 (85.5)<sup>b</sup> | 1,464 (58.2)<sup>b</sup> | 1,002 (42.2)<sup>b</sup> | 920 (39.9)<sup>b</sup> |
| Both at work and remote | 41 (3.0) | 75 (3.0) | 66 (2.8) | 52 (2.3) |
| Remote | 46 (3.4) | 215 (8.5) | 217 (9.2) | 202 (8.8) |
| Did not work | 110 (8.1) | 761 (30.3) | 1,087 (45.8) | 1,130 (49.0) |
| **COVID-19 pandemic period** | (n = 2,188) | (n = 2,509) | (n = 2,418) | (n = 2,382) |
| At work | 1,676 (76.6) | 1,239 (49.4) | 644 (26.6) | 561 (23.5) |
| Both at work and remote | 73 (3.3) | 83 (3.3) | 42 (1.8) | 43 (1.8) |
| Remote | 251 (11.5) | 380 (15.1) | 474 (19.6) | 440 (18.5) |
| Did not work | 188 (8.6) | 807 (32.2) | 1,258 (52.0) | 1,338 (56.2) |

<sup>a</sup>Persons with only work from home experience before illness onset (560, 676, 689, and 677 for day before, day 1, day 2, and day 3, respectively) and those with missing work location (41, 74, 55, and 50 for day before, day 1, day 2, and day 3, respectively) were excluded.
<sup>b</sup>p<0.001 (comparison of pre-pandemic to pandemic period for specified day).

Participants with hybrid experience were less likely to work onsite than persons with in-person only experience on the day before illness and during the first 3 days of illness (Table 3). The magnitude of the effect was more pronounced for the pandemic period than the pre-pandemic influenza seasons. For example, on the third day of illness, aOR for hybrid vs. in-person only experience was 0.69 (95% CI 0.54–0.87) for the pre-pandemic period compared to 0.38 (95% CI 0.29–0.49) for the pandemic period (p<0.001 for the work experience*study period interaction term). Conversely, participants were less likely to work onsite during the COVID-19 pandemic period than the pre-pandemic influenza seasons, with the magnitude of the effect being more pronounced among persons with hybrid experience than among persons with in-person only experience (e.g., for the third day of illness, aOR for pandemic vs. pre-pandemic period was 0.32 among persons with hybrid experience compared to 0.59 among persons with in-person only experience) (Table 3). Persons with hybrid experience were more likely to work remotely during the pandemic period than the pre-pandemic period (Appendix Table 5). In contrast, persons with in-person only experience were more likely to abstain from work during the pandemic period than the pre-pandemic period. Restricting the regression models to non-healthcare personnel or to the five sites that contributed data for all four study years showed similar findings (Appendix Tables 6–7). The findings were also similar when the analysis was restricted to the five sites with higher survey completion rates (Appendix Table 8).

**Table 3.**
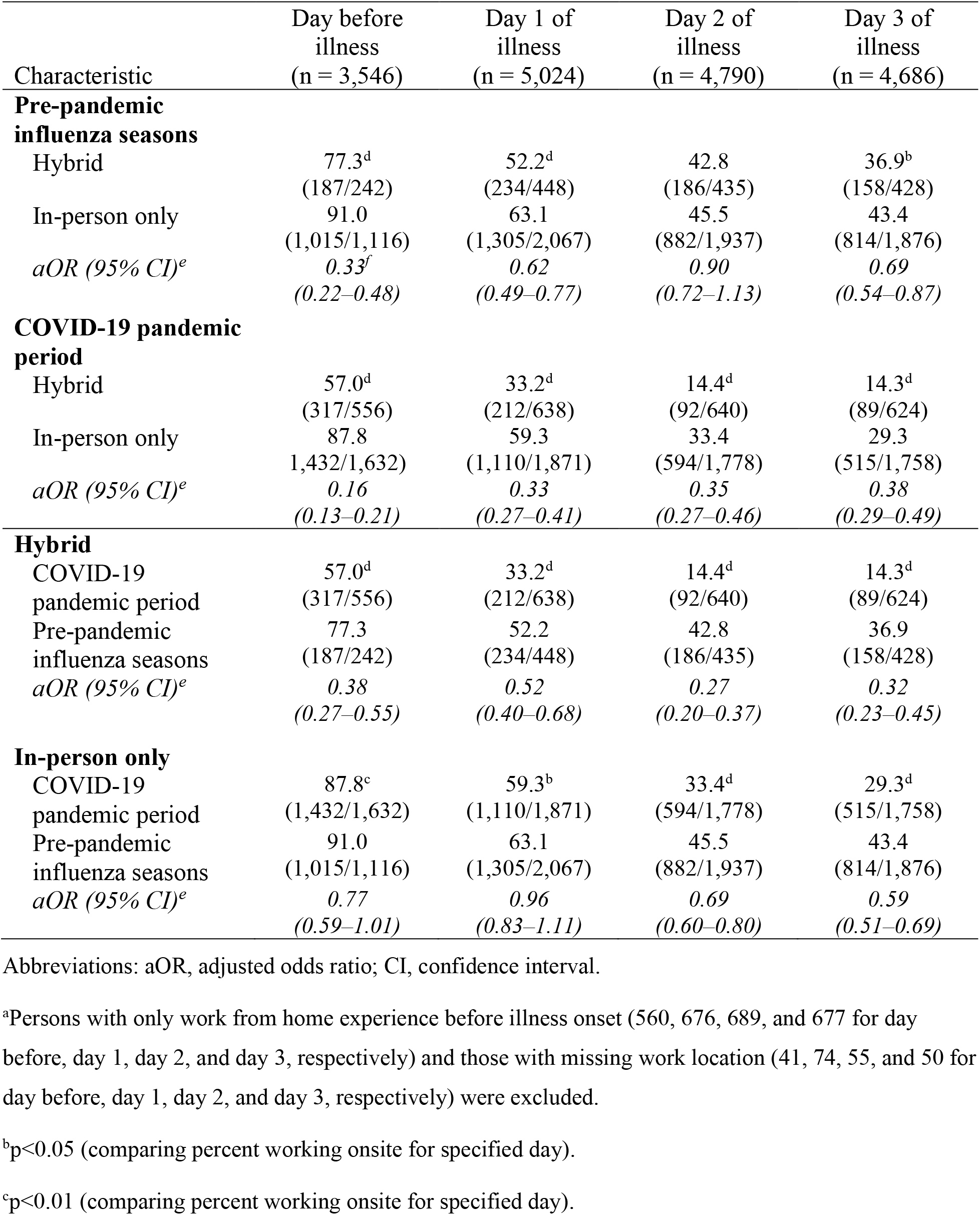
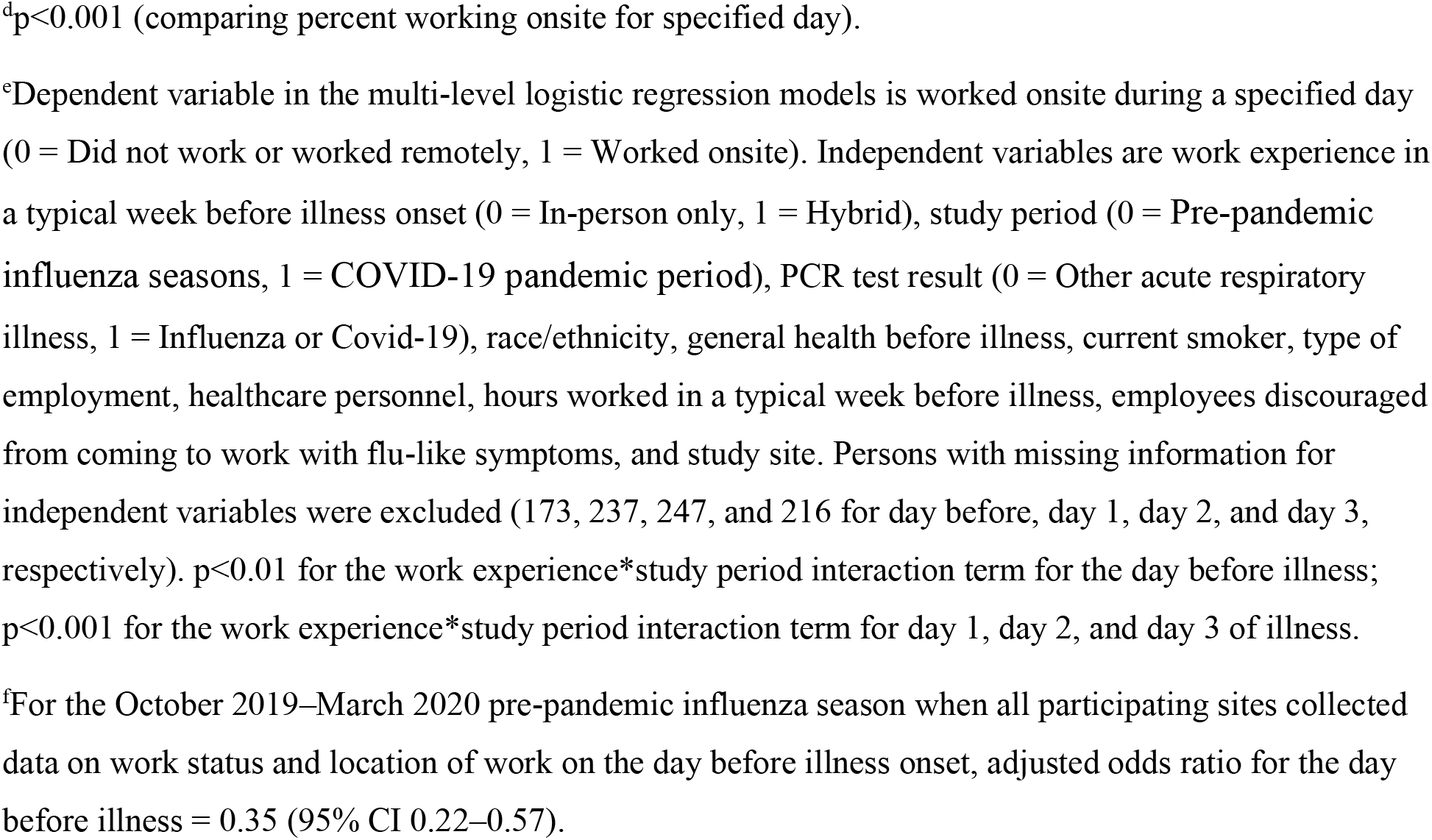
Likelihood of working onsite among adults with influenza, COVID-19, or other acute respiratory illness who were scheduled to work, by prior experience working from home, United States, 2018–2022^a^.

| Characteristic | Day before illness<br>(n = 3,546) | Day 1 of illness<br>(n = 5,024) | Day 2 of illness<br>(n = 4,790) | Day 3 of illness<br>(n = 4,686) |
| --- | --- | --- | --- | --- |
| **Pre-pandemic influenza seasons** |  |  |  |  |
| Hybrid | 77.3<sup>d</sup><br>(187/242) | 52.2<sup>d</sup><br>(234/448) | 42.8<br>(186/435) | 36.9<sup>b</sup><br>(158/428) |
| In-person only | 91.0<br>(1,015/1,116) | 63.1<br>(1,305/2,067) | 45.5<br>(882/1,937) | 43.4<br>(814/1,876) |
| *aOR (95% CI)<sup>e</sup>* | *0.33<sup>f</sup>*<br>(0.22–0.48) | *0.62*<br>(0.49–0.77) | *0.90*<br>(0.72–1.13) | *0.69*<br>(0.54–0.87) |
| **COVID-19 pandemic period** |  |  |  |  |
| Hybrid | 57.0<sup>d</sup><br>(317/556) | 33.2<sup>d</sup><br>(212/638) | 14.4<sup>d</sup><br>(92/640) | 14.3<sup>d</sup><br>(89/624) |
| In-person only | 87.8<br>(1,432/1,632) | 59.3<br>(1,110/1,871) | 33.4<br>(594/1,778) | 29.3<br>(515/1,758) |
| *aOR (95% CI)<sup>e</sup>* | *0.16*<br>(0.13–0.21) | *0.33*<br>(0.27–0.41) | *0.35*<br>(0.27–0.46) | *0.38*<br>(0.29–0.49) |
| **Hybrid** |  |  |  |  |
| COVID-19 pandemic period | 57.0<sup>d</sup><br>(317/556) | 33.2<sup>d</sup><br>(212/638) | 14.4<sup>d</sup><br>(92/640) | 14.3<sup>d</sup><br>(89/624) |
| Pre-pandemic influenza seasons | 77.3<br>(187/242) | 52.2<br>(234/448) | 42.8<br>(186/435) | 36.9<br>(158/428) |
| *aOR (95% CI)<sup>e</sup>* | *0.38*<br>(0.27–0.55) | *0.52*<br>(0.40–0.68) | *0.27*<br>(0.20–0.37) | *0.32*<br>(0.23–0.45) |
| **In-person only** |  |  |  |  |
| COVID-19 pandemic period | 87.8<sup>c</sup><br>(1,432/1,632) | 59.3<sup>b</sup><br>(1,110/1,871) | 33.4<sup>d</sup><br>(594/1,778) | 29.3<sup>d</sup><br>(515/1,758) |
| Pre-pandemic influenza seasons | 91.0<br>(1,015/1,116) | 63.1<br>(1,305/2,067) | 45.5<br>(882/1,937) | 43.4<br>(814/1,876) |
| *aOR (95% CI)<sup>e</sup>* | *0.77*<br>(0.59–1.01) | *0.96*<br>(0.83–1.11) | *0.69*<br>(0.60–0.80) | *0.59*<br>(0.51–0.69) |

Abbreviations: aOR, adjusted odds ratio; CI, confidence interval.
<sup>a</sup>Persons with only work from home experience before illness onset (560, 676, 689, and 677 for day before, day 1, day 2, and day 3, respectively) and those with missing work location (41, 74, 55, and 50 for day before, day 1, day 2, and day 3, respectively) were excluded.
<sup>b</sup>p<0.05 (comparing percent working onsite for specified day).
<sup>c</sup>p<0.01 (comparing percent working onsite for specified day).
<sup>d</sup> $p < 0.001$ (comparing percent working onsite for specified day).
<sup>e</sup>Dependent variable in the multi-level logistic regression models is worked onsite during a specified day (0 = Did not work or worked remotely, 1 = Worked onsite). Independent variables are work experience in a typical week before illness onset (0 = In-person only, 1 = Hybrid), study period (0 = Pre-pandemic influenza seasons, 1 = COVID-19 pandemic period), PCR test result (0 = Other acute respiratory illness, 1 = Influenza or Covid-19), race/ethnicity, general health before illness, current smoker, type of employment, healthcare personnel, hours worked in a typical week before illness, employees discouraged from coming to work with flu-like symptoms, and study site. Persons with missing information for independent variables were excluded (173, 237, 247, and 216 for day before, day 1, day 2, and day 3, respectively). $p < 0.01$ for the work experience\*study period interaction term for the day before illness; $p < 0.001$ for the work experience\*study period interaction term for day 1, day 2, and day 3 of illness.
<sup>f</sup>For the October 2019–March 2020 pre-pandemic influenza season when all participating sites collected data on work status and location of work on the day before illness onset, adjusted odds ratio for the day before illness = 0.35 (95% CI 0.22–0.57).

Stratifying the analysis by PCR test result showed that the proportion who did not work during illness was greater for persons with influenza than that for other ARI (Appendix Table 9). The proportion who did not work was also greater for persons with COVID-19 than that for other ARI. For example, on the third day of illness during the pre-pandemic influenza seasons, the proportion who did not work was 64.4% for persons with influenza and 40.3% for persons with other ARI (p<0.001). During the pandemic period, the proportion who did not work on the third day of illness was 66.7% for persons with COVID-19 and 48.3% for persons with other ARI (p<0.001).

For the pre-pandemic influenza seasons, persons with influenza were significantly less likely to work onsite than persons with other ARI on the second (aOR 0.51, 95% CI 0.43–0.61) and third days of illness (aOR 0.39, 95% CI 0.32–0.47) (Table 4). For the pandemic period, participants with COVID-19 were significantly less likely to work onsite than persons with other ARI on the second (aOR 0.59, 95% CI 0.49–0.73) and third days of illness (aOR 0.31, 95% CI 0.25–0.39).

**Table 4.**
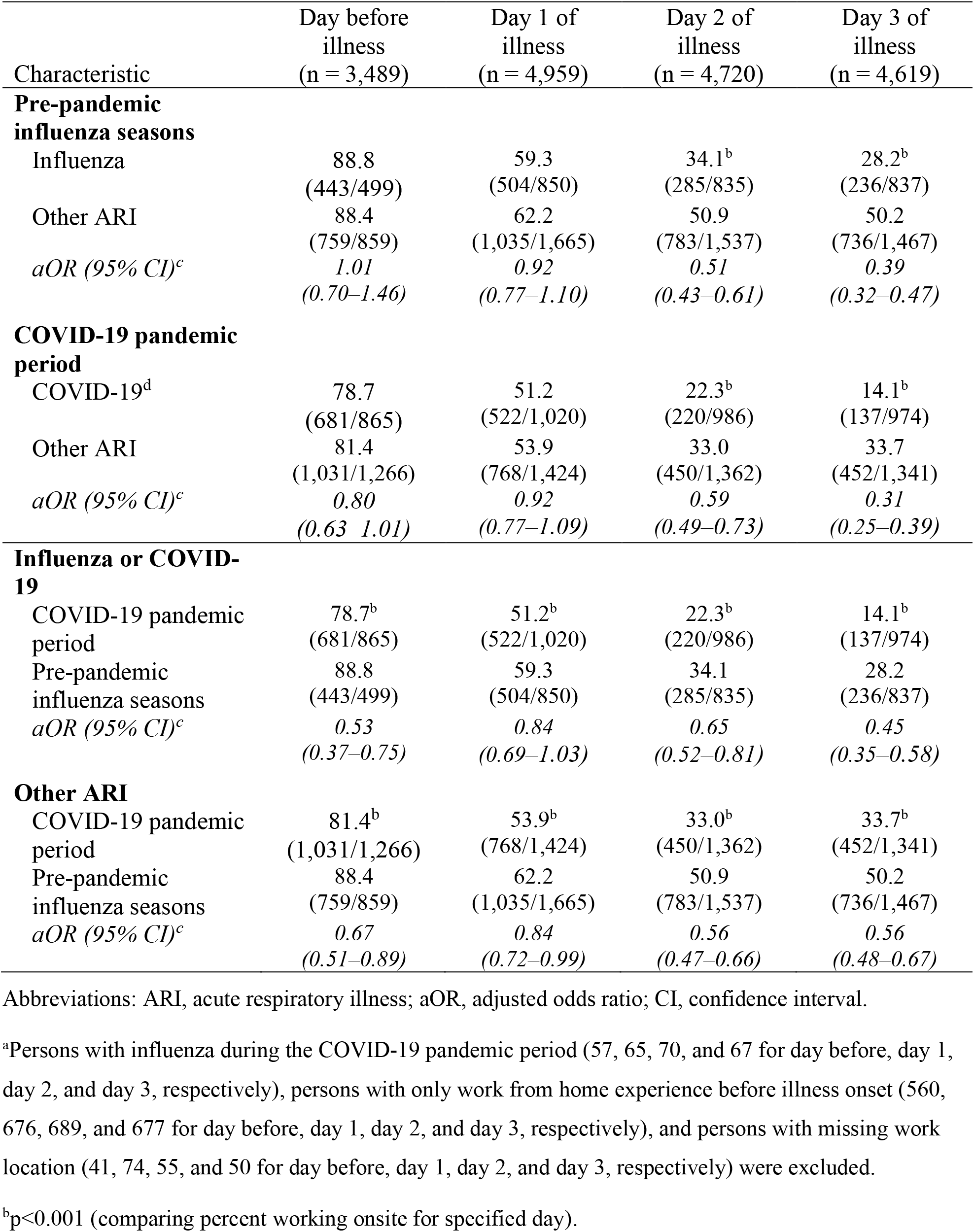
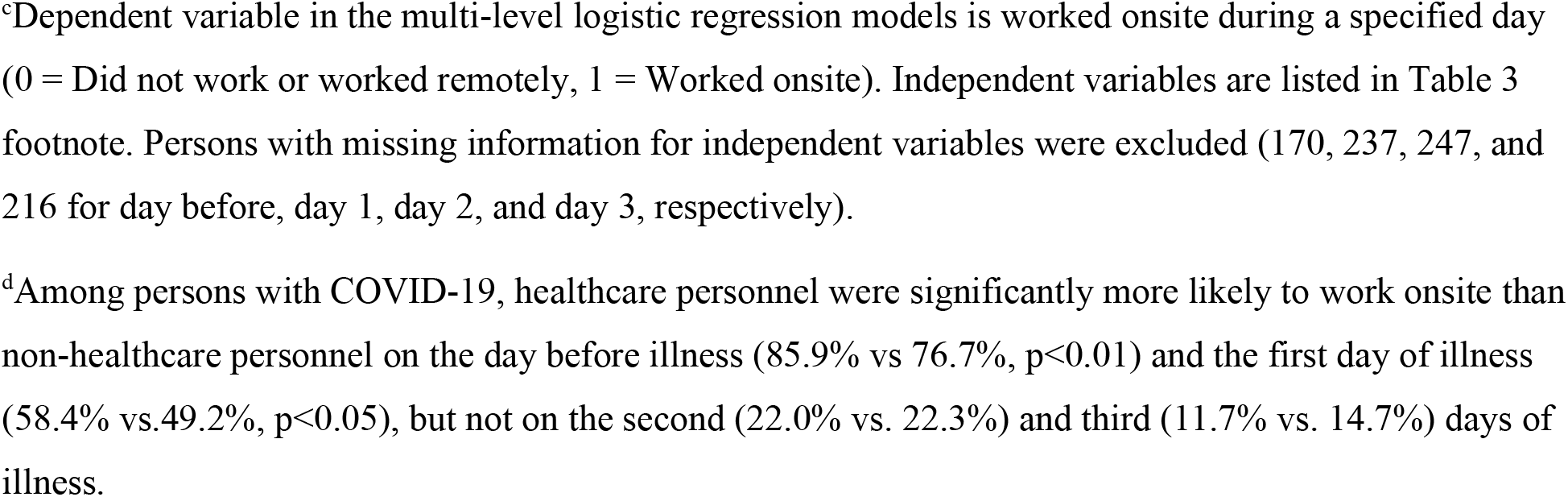
Likelihood of working onsite among adults who were scheduled to work, by PCR test result, United States, 2018–2022^a^.

| Characteristic | Day before illness<br>(n = 3,489) | Day 1 of illness<br>(n = 4,959) | Day 2 of illness<br>(n = 4,720) | Day 3 of illness<br>(n = 4,619) |
| --- | --- | --- | --- | --- |
| **Pre-pandemic influenza seasons** |  |  |  |  |
| Influenza | 88.8<br>(443/499) | 59.3<br>(504/850) | 34.1<sup>b</sup><br>(285/835) | 28.2<sup>b</sup><br>(236/837) |
| Other ARI | 88.4<br>(759/859) | 62.2<br>(1,035/1,665) | 50.9<br>(783/1,537) | 50.2<br>(736/1,467) |
| *aOR (95% CI)<sup>c</sup>* | *1.01*<br>(0.70–1.46) | *0.92*<br>(0.77–1.10) | *0.51*<br>(0.43–0.61) | *0.39*<br>(0.32–0.47) |
| **COVID-19 pandemic period** |  |  |  |  |
| COVID-19<sup>d</sup> | 78.7<br>(681/865) | 51.2<br>(522/1,020) | 22.3<sup>b</sup><br>(220/986) | 14.1<sup>b</sup><br>(137/974) |
| Other ARI | 81.4<br>(1,031/1,266) | 53.9<br>(768/1,424) | 33.0<br>(450/1,362) | 33.7<br>(452/1,341) |
| *aOR (95% CI)<sup>c</sup>* | *0.80*<br>(0.63–1.01) | *0.92*<br>(0.77–1.09) | *0.59*<br>(0.49–0.73) | *0.31*<br>(0.25–0.39) |
| **Influenza or COVID-19** |  |  |  |  |
| COVID-19 pandemic period | 78.7<sup>b</sup><br>(681/865) | 51.2<sup>b</sup><br>(522/1,020) | 22.3<sup>b</sup><br>(220/986) | 14.1<sup>b</sup><br>(137/974) |
| Pre-pandemic influenza seasons | 88.8<br>(443/499) | 59.3<br>(504/850) | 34.1<br>(285/835) | 28.2<br>(236/837) |
| *aOR (95% CI)<sup>c</sup>* | *0.53*<br>(0.37–0.75) | *0.84*<br>(0.69–1.03) | *0.65*<br>(0.52–0.81) | *0.45*<br>(0.35–0.58) |
| **Other ARI** |  |  |  |  |
| COVID-19 pandemic period | 81.4<sup>b</sup><br>(1,031/1,266) | 53.9<sup>b</sup><br>(768/1,424) | 33.0<sup>b</sup><br>(450/1,362) | 33.7<sup>b</sup><br>(452/1,341) |
| Pre-pandemic influenza seasons | 88.4<br>(759/859) | 62.2<br>(1,035/1,665) | 50.9<br>(783/1,537) | 50.2<br>(736/1,467) |
| *aOR (95% CI)<sup>c</sup>* | *0.67*<br>(0.51–0.89) | *0.84*<br>(0.72–0.99) | *0.56*<br>(0.47–0.66) | *0.56*<br>(0.48–0.67) |

Abbreviations: ARI, acute respiratory illness; aOR, adjusted odds ratio; CI, confidence interval.
<sup>a</sup>Persons with influenza during the COVID-19 pandemic period (57, 65, 70, and 67 for day before, day 1, day 2, and day 3, respectively), persons with only work from home experience before illness onset (560, 676, 689, and 677 for day before, day 1, day 2, and day 3, respectively), and persons with missing work location (41, 74, 55, and 50 for day before, day 1, day 2, and day 3, respectively) were excluded.
<sup>b</sup>p<0.001 (comparing percent working onsite for specified day).
<sup>c</sup>Dependent variable in the multi-level logistic regression models is worked onsite during a specified day (0 = Did not work or worked remotely, 1 = Worked onsite). Independent variables are listed in Table 3 footnote. Persons with missing information for independent variables were excluded (170, 237, 247, and 216 for day before, day 1, day 2, and day 3, respectively).
<sup>d</sup>Among persons with COVID-19, healthcare personnel were significantly more likely to work onsite than non-healthcare personnel on the day before illness (85.9% vs 76.7%, $p<0.01$ ) and the first day of illness (58.4% vs. 49.2%, $p<0.05$ ), but not on the second (22.0% vs. 22.3%) and third (11.7% vs. 14.7%) days of illness.

Among persons with COVID-19, a substantial proportion worked onsite during illness: 51.2% on day 1; 22.3% on day 2; 14.1% on day 3 (Table 4). COVID-19 positive PCR test results were available for 1.3% (12/940) by the first day of COVID-19 illness, 10.7% (97/910) by the second day, and 23.5% (211/899) by the third day (Table 5). Persons for whom a positive COVID-19 PCR test result was available by the second day of illness were significantly less likely to work onsite on that day than those whose positive PCR result was available after the second day of illness (5.2% vs. 25.0%, p<0.001) (Table 5). Persons for whom a positive PCR test result was available by the third day of illness were significantly less likely to work onsite on that day than those whose positive PCR result was available after the third day of illness (4.7% vs. 17.2%, p<0.001). Among persons for whom positive PCR test results were available after the second or third day of illness, excluding persons with COVID-19 positive *at-home* test results by the second or third day of illness resulted in a slight increase in the percent who worked onsite (Appendix Table 10).

**Table 5.** Likelihood of working onsite among adults with COVID-19 illness who were scheduled to work, by day when COVID-19 positive PCR test result was available, United States, October 2020–June 2022^a^.

| Characteristic | Worked onsite, % |
| --- | --- |
| Scheduled to work on day 1 of COVID-19 illness<sup>b</sup> |  |
| COVID-19 positive PCR result available on day 1 of illness | 50.0 (6/12) |
| COVID-19 positive PCR result available after day 1 of illness | 52.1 (483/928) |
| Scheduled to work on day 2 of COVID-19 illness<sup>c</sup> |  |
| COVID-19 positive PCR result available on day 1 or 2 of illness | 5.2 (5/97)<sup>e</sup> |
| COVID-19 positive PCR result available after day 2 of illness | 25.0 (203/813) |
| Scheduled to work on day 3 of COVID-19 illness<sup>d</sup> |  |
| COVID-19 positive PCR result available on day 1, 2, or 3 of illness | 4.7 (10/211)<sup>e</sup> |
| COVID-19 positive PCR result available after day 3 of illness | 17.2 (118/688) |

Values represent percent worked onsite (at work + both at work and remotely / scheduled to work).
Analysis is restricted to persons with COVID-19 shown in Table 4.
<sup>a</sup>Day of illness when COVID-19 positive result was available was computed by comparing the date of illness onset with the date that COVID-19 positive PCR test result was available.
<sup>b</sup>Unknown when COVID-19 positive PCR result was available = 80 persons.
<sup>c</sup>Unknown when COVID-19 positive PCR result was available = 76 persons.
<sup>d</sup>Unknown when COVID-19 positive PCR result was available = 75 persons.
<sup>e</sup>p<0.001.

## Discussion

For both the pre-pandemic and pandemic periods, adults with WFH or hybrid experience were more able to work during the first 3 days of illness compared to those with in-person only experience. It is important to note, however, that persons with hybrid experience were significantly less likely to work onsite than those with only in-person experience on the day before illness and during the first 3 days of illness. The magnitude of the effect of hybrid vs. in-person experience on working onsite while ill was more pronounced for the pandemic period than the pre-pandemic period. Persons with influenza or COVID-19 were significantly less likely to work onsite than persons with other ARI on the second and third days of illness. For persons for whom a positive COVID-19 PCR test result was available by the second or third day of illness, few reported working onsite.

Persons with prior WFH and hybrid experience were significantly more likely to work during illness than those with only in-person experience, enabling a greater level of continuity of work during illness without risk of infecting others at the workplace. A greater likelihood of working among persons with hybrid experience than among those with only in-person experience has been reported in studies conducted during the 2017–2018 influenza season and during the early part of the COVID-19 pandemic (March–November 2020) (10, 17). WFH or hybrid experience before illness can enable persons, who are well enough, to work remotely instead of taking a sick day. It is possible that persons without experience working from home were more likely to be in occupations where WFH or hybrid work is less feasible, and workers are less likely to have the option or incentive to work remotely. These occupations might include hospitality and leisure, transportation, utilities, construction, production, and agriculture (18, 19). Employers were required to provide paid sick leave to workers with COVID-19 during the pandemic (20). It is unlikely that persons in occupations less amenable to working from home worked less because they received paid sick leave after testing SARS-CoV-2 positive than persons with hybrid experience. This pattern was also observed before the pandemic.

Persons with hybrid experience before illness onset were less likely to work onsite on the day before illness and during the first 3 days of illness than persons with only in-person experience, thus reducing the likelihood of workplace exposures to respiratory viruses. A study conducted during the 2017–2018 influenza season reported that persons with hybrid experience before illness onset were less likely to work onsite during the first 3 days of illness than persons with in-person only experience, but the study did not examine the likelihood of working onsite on the day before illness (17). A study conducted during the early part of the COVID-19 pandemic found that persons with hybrid experience were less likely to work onsite during illness than persons with in-person experience, but it is unclear whether persons worked onsite early during their illness when infectiousness is higher (10). From this study, it is clear that persons with hybrid work experience are less likely to work onsite in general. The finding that the effect of hybrid work experience vs. in-person only experience was more pronounced during the pandemic period than the pre-pandemic period may be because of the greater prevalence of telework during the pandemic regardless of illness (3, 4). The intense public health messaging to stay home when ill during the COVID-19 pandemic, employer policies that discouraged or prohibited employees with symptoms of ARI to work onsite during the pandemic, and the provision of flexible paid leave for persons with COVID-19 illness may have contributed to the greater effect during the pandemic period (5, 20).

This study shows that persons with laboratory-confirmed influenza or COVID-19 were significantly less likely to work onsite during the second and third days of illness than persons with other ARI. Previous studies have reported similar findings, but the studies did not assess the likelihood of working onsite on each of the first 3 days of illness (10, 17). These findings may be attributed to more severe illness in persons with influenza or COVID-19 (15). The finding that the likelihood of working onsite was similar on the first day of illness, as well as the greater likelihood of working onsite on the first day of illness compared to the second or third day of illness, may be because illness might have begun when participants were already at work. For persons with COVID-19 illness, availability of positive PCR test results by the second or third day of illness might have reduced the likelihood of working onsite for several reasons, including concern for co-workers, being advised to isolate by case investigators, employers discouraging or prohibiting persons with COVID-19 from entering the worksite, and employers providing flexible sick leave. However, COVID-19 positive PCR test results were available for a small proportion of persons during the first 3 days of illness because of the lag between illness onset and seeking medical care. At-home rapid COVID-19 tests may facilitate early testing for persons with symptoms of ARI. The use of at-home tests among persons with COVID-19-like illness in the United States increased from 6% in August 23–December 11, 2021 to 20% in December 19, 2021–March 12, 2022 (21).

A strength of our study is that about 8,000 persons participated over 4 years that encompassed both the pre-pandemic and pandemic periods. We obtained respiratory specimens that enabled laboratory confirmation of influenza and SARS-CoV-2. We assessed work attendance during the pre-symptomatic phase when persons can be infectious and during the first 3 days of illness when infectiousness is greatest. Our study has some limitations. First, a sizeable proportion of persons (39%) did not complete the follow-up survey. However, the findings were similar when the analysis was restricted to the five sites with higher survey completion rates. Second, we assessed the proportion who worked during the first 3 days of illness as an indicator of maintenance of workflow. We did not assess how illness may have diminished productivity. Third, we assessed work attendance among persons with medically attended ARIs. The findings may not be generalizable to persons who were asymptomatic or who did not seek medical care. Future research should ascertain productivity in persons who work during influenza or COVID-19 illness and assess the likelihood of working onsite among persons with ARI who do not seek medical care. Research is also needed on how occupation and workplace policies affect work attendance during illness.

Working-age adults continue to be at risk of severe COVID-19 (22). Our study findings show that hybrid experience before illness onset may give workers the option to continue working but also reduce working onsite early during their illness when infectiousness is higher. Persons with hybrid work experience were less likely to work onsite during both the pre-symptomatic and symptomatic phases of illness. Work from home policies, for occupations where feasible, may reduce the likelihood of workplace exposures to influenza and SARS-CoV-2 viruses.

## Data Availability

The data produced in the study are not available online.

## Appendix

**Appendix Table 1.** Follow-up survey questions, 2018–2022^a^.

| Question | Values |
| --- | --- |
| Have you fully or mostly recovered from this illness? | Yes: Date: __-__-____ (mm/dd/yyyy)<br>I have not fully or mostly recovered from this illness<br>Don't know<br>Refused |
| Are you currently employed (work for pay or profit)? | I work for an employer<br>I am self-employed or own my own business → [Survey is complete]<br>No → [Survey is complete]<br>Refused → [Survey is complete] |
| How many hours are you <u>expected</u> to work in a typical 7-day week?<sup>b</sup><br>(If it varies, estimate the average) | Number of hours |
| Of those expected hours, how many hours in a week do you usually work from home (telework, telecommute, or remote work)?<sup>c</sup><br>(Enter “0” if none) | Number of hours |
| Are you salaried or are you paid hourly? [“Salaried” means you’re paid the same amount each week or month no matter how many hours you work. “Hourly” means that you’re paid a different amount each week or month depending on how many hours you work.] | Salaried<br>Paid hourly<br>Other such as commission only<br>Don't know<br>Refused |
| Do you work in a healthcare setting with direct patient contact?<sup>d</sup> | Yes<br>No<br>Don't know |
|  | Refused |
| Please think about the first three days of your illness.<br>The first day of illness being the day your symptoms started [DAY/DATE OF ONSET] and the third day of illness being [DATE OF ONSET + 2]. |  |
| On the <u>first</u> day of illness [DATE OF ONSET]: |  |
| How many hours were you scheduled to work? | Number of hours |
| How many hours did you work? | Number of hours |
| Where did you work? | At work<br>Remotely<br>Both at work and remotely<br>Don't know<br>Refused |
| On the <u>second</u> day of illness [DATE OF ONSET+1]: |  |
| How many hours were you scheduled to work? | Number of hours |
| How many hours did you work? | Number of hours |
| Where did you work? | At work<br>Remotely<br>Both at work and remotely<br>Don't know<br>Refused |
| On the <u>third</u> day of illness [DATE OF ONSET+2]: |  |
| How many hours were you scheduled to work? | Number of hours |
| How many hours did you work? | Number of hours |
| Where did you work? | At work<br>Remotely<br>Both at work and remotely<br>Don't know<br>Refused |
| On the day <u>before</u> your illness began [DATE OF ONSET-1]:<sup>e</sup> |  |
| How many hours were you scheduled to work? | Number of hours |
| How many hours did you work? | Number of hours |
| Where did you work? | At work<br>Remotely<br>Both at work and remotely |
|  | Don't know<br>Refused |
| Please select your level of agreement with the following statement about your place of work:<br>- Employees are discouraged from coming to work when they have flu-like symptoms | Strongly agree<br>Agree<br>Neither agree nor disagree<br>Disagree<br>Strongly disagree |

<sup>a</sup>If a person worked multiple jobs, the person was asked to think about the job that they considered as their primary job when answering the questions.
<sup>b</sup>For the COVID-19-dominated period, the wording was “During the month before illness, how many hours were you expected to work in a week?”.
<sup>c</sup>For the COVID-19-dominated period, the wording was: “Of those expected hours, how many hours in a week did you usually work from home (telework, telecommute, or remote work)?
<sup>d</sup>For the COVID-19-dominated period, the question was asked at enrollment: “Did you work in a healthcare setting and have close contact with patients during the 14 days before your illness began? Close contact means being within 6 feet of a patient.”
<sup>e</sup>This was an optional question for the period November 2018–May 2019.

**Appendix Table 2.** Data collection by study sites, 2018–2022^a^.

| Site | Nov 2018–May 2019 |  | Oct 2019–Mar 2020 |  | Oct 2020–Sep 2021 |  | Oct 2021–Jun 2022 |  |
| --- | --- | --- | --- | --- | --- | --- | --- | --- |
|  | D0 | D1, D2, D3 | D0 | D1, D2, D3 | D0 | D1, D2, D3 | D0 | D1, D2, D3 |
| MI |  | ✓ | ✓ | ✓ | ✓ | ✓ | ✓ | ✓ |
| PA | ✓ | ✓ | ✓ | ✓ | ✓ | ✓ | ✓ | ✓ |
| TX |  | ✓ | ✓ | ✓ | ✓ | ✓ | ✓ | ✓ |
| WA<sup>b</sup> |  | ✓ | ✓ | ✓ | ✓ | ✓ | ✓ | ✓ |
| WI<sup>b</sup> |  | ✓ | ✓ | ✓ | ✓ | ✓ | ✓ | ✓ |
| CA |  |  |  |  |  |  | ✓ | ✓ |
| TN |  |  |  |  |  |  | ✓ | ✓ |

Abbreviations: D0, day before illness; D1, first day of illness; D2, second day of illness; D3, third day of illness; MI, Michigan; PA, Pennsylvania; TX, Texas; WA, Washington; WI, Wisconsin; CA, California; TN, Tennessee.
<sup>a</sup>Check mark indicates that a site collected data for the specified period and day of illness.
<sup>b</sup>For persons with only work from home experience before illness onset, the Washington and Wisconsin sites did not collect data on work status (hours scheduled to work, hours worked) and location of work for the period November 2018 to September 2021.

**Appendix Table 3.** Characteristics of adults with COVID-19, influenza, or other acute respiratory illness, United States, 2018–2022.

| Characteristic | Work experience in a typical week before illness onset |  |  |
| --- | --- | --- | --- |
|  | Work from home only (n = 1,139) | Hybrid (n = 1,503) | In-person only (n = 5,490) |
| Study period<sup>c</sup> |  |  |  |
| Pre-pandemic influenza seasons | 126 (11.1) | 628 (41.8) | 2,853 (52.0) |
| COVID-19 pandemic period | 1,013 (88.9) | 875 (58.2) | 2,637 (48.0) |
| Median hours worked in a typical week before illness (IQR)<sup>c</sup> | 40 (40, 40) | 40 (40, 45) | 40 (40, 40) |
| Median hours usually worked from home in a typical week before illness (IQR) |  |  |  |
| Pre-pandemic influenza seasons | 40 (40, 45)<sup>a</sup> | 8 (5, 15)<sup>c</sup> | 0 (0, 0) |
| COVID-19 pandemic period | 40 (40, 40) | 16 (8, 25) | 0 (0, 0) |
| Type of employment<sup>c</sup> |  |  |  |
| Hourly | 281 (29.9) | 325 (21.8) | 3,618 (66.6) |
| Salaried or other | 658 (70.1) | 1,166 (78.2) | 1,816 (33.4) |
| Healthcare personnel<sup>c</sup> |  |  |  |
| Yes | 78 (7.1) | 231 (15.5) | 1,379 (25.4) |
| No | 1,017 (92.9) | 1,264 (84.5) | 4,055 (74.6) |
| Employees discouraged from coming to work with flu-like symptoms<sup>c</sup> |  |  |  |
| Agree | 805 (88.6) | 1,293 (86.8) | 4,175 (76.7) |
| Not agree | 104 (11.4) | 196 (13.2) | 1,266 (23.3) |
| Median age, y (IQR)<sup>c</sup> | 40 (33, 51) | 40 (33, 51) | 39 (30, 51) |
| Sex<sup>b</sup> |  |  |  |
| Female | 781 (68.8) | 939 (62.5) | 3,651 (66.5) |
| Male | 354 (31.2) | 564 (37.5) | 1,835 (33.5) |
| Race/ethnicity<sup>b</sup> |  |  |  |
| White, non-Hispanic | 846 (74.8) | 1,142 (76.5) | 4,222 (77.4) |
| Black, non-Hispanic | 63 (5.6) | 68 (4.5) | 335 (6.1) |
| Other, non-Hispanic | 122 (10.8) | 152 (10.2) | 422 (7.7) |
| Hispanic, any race | 100 (8.8) | 131 (8.8) | 479 (8.8) |
| Education<sup>c</sup> |  |  |  |
| High school or less | 90 (7.9) | 50 (3.3) | 1,073 (19.6) |
| Some college | 236 (20.8) | 258 (17.2) | 2,021 (36.9) |
| Bachelor's degree | 471 (41.4) | 595 (39.7) | 1,503 (27.4) |
|  | Work from home only (n = 1,139) | Hybrid (n = 1,503) | In-person only (n = 5,490) |
| Advanced degree | 340 (29.9) | 597 (39.8) | 883 (16.1) |
| General health before illness<sup>b</sup> |  |  |  |
| Excellent | 203 (18.4) | 316 (21.4) | 991 (18.3) |
| Very good | 473 (42.9) | 652 (44.1) | 2,244 (41.5) |
| Good | 347 (31.5) | 418 (28.3) | 1,743 (32.2) |
| Fair/poor | 79 (7.2) | 91 (6.2) | 434 (8.0) |
| Current smoker<sup>c</sup> |  |  |  |
| Yes | 57 (5.1) | 99 (6.7) | 647 (11.9) |
| No | 1,059 (94.9) | 1,378 (93.3) | 4,777 (88.1) |
| Children <12 y of age in household<sup>a</sup> |  |  |  |
| 0 | 745 (65.7) | 1,002 (67.0) | 3,812 (69.6) |
| 1 | 199 (17.6) | 252 (16.8) | 810 (14.8) |
| ≥2 | 190 (16.7) | 242 (16.2) | 856 (15.6) |
| Fully or mostly recovered from illness at follow-up |  |  |  |
| Yes | 834 (74.9) | 1,079 (73.1) | 3,879 (72.1) |
| No | 279 (25.1) | 398 (26.9) | 1,499 (27.9) |
| Among persons fully or mostly recovered from illness, median duration of illness, days (IQR)<sup>b</sup> | 9 (7, 12) | 9 (7,12) | 9 (7, 11) |
| Median days from illness onset to enrollment (IQR)<sup>c</sup> | 4 (2, 7) | 3 (2, 6) | 3 (2, 5) |
| Median days from illness onset to follow-up (IQR)<sup>c</sup> | 13 (10, 17) | 12 (10, 15) | 12 (10, 14) |
| Site<sup>c</sup> |  |  |  |
| Michigan | 91 (8.0) | 169 (11.2) | 405 (7.4) |
| Pennsylvania | 323 (28.4) | 410 (27.3) | 1,352 (24.6) |
| Texas | 59 (5.2) | 91 (6.1) | 604 (11.0) |
| Washington | 238 (20.9) | 346 (23.0) | 1,143 (20.8) |
| Wisconsin | 172 (15.1) | 182 (12.1) | 1,354 (24.7) |
| California | 136 (11.9) | 169 (11.2) | 299 (5.4) |
| Tennessee | 120 (10.5) | 136 (9.1) | 333 (6.1) |

Abbreviation: IQR, interquartile range.
Values are no. (column %) except as indicated. Numbers may not sum to n because of missing data.
<sup>a</sup>p<0.05. <sup>b</sup>p<0.01. <sup>c</sup>p<0.001.

**Appendix Table 4.** Proportion of persons with COVID-19, influenza, or other acute respiratory illness who were scheduled to work, United States, 2018–2022^a^.

| Work experience in a typical week before illness onset | Day before illness | Day 1 of illness | Day 2 of illness | Day 3 of illness |
| --- | --- | --- | --- | --- |
|  | Scheduled to work, %<sup>b</sup> |  |  |  |
| Work from home only | 64.6<br>(560/867) | 77.1<br>(676/877) | 77.5<br>(689/889)<sup>c</sup> | 76.0<br>(677/891)<sup>c</sup> |
| Hybrid | 66.3<br>(810/1,221) | 76.3<br>(1,109/1,454) | 74.5<br>(1,092/1,465) | 73.5<br>(1,071/1,458) |
| In-person only | 66.0<br>(2,777/4,210) | 75.4<br>(3,989/5,290) | 70.5<br>(3,753/5,326) | 69.2<br>(3,665/5,300) |
| TOTAL | 65.9<br>(4147/6,298) | 75.8<br>(5,774/7,621) | 72.1<br>(5,534/7,680) | 70.8<br>(5,413/7,649) |

<sup>a</sup>Persons were excluded if data on hours scheduled to work or hours worked were missing: 1,834 on the day before illness, 511 on day 1, 452 on day 2, and 483 on day 3. Among the 1,834 persons who were excluded on the day before illness, 1,423 were excluded because the Michigan, Texas, Washington, and Wisconsin sites did not collect data on work status (hours scheduled to work, hours worked) for the day before illness during November 2018–May 2019.
<sup>b</sup>Numerator represents number scheduled to work and denominator represents sum of number scheduled to work and number not scheduled to work. Among persons who were scheduled to work, the median (IQR) hours scheduled to work was 8 (8, 8) for the day before illness and for each day of illness.
<sup>c</sup> $p < 0.001$ (comparison of the three work experience categories for specified day).

**Appendix Table 5.**
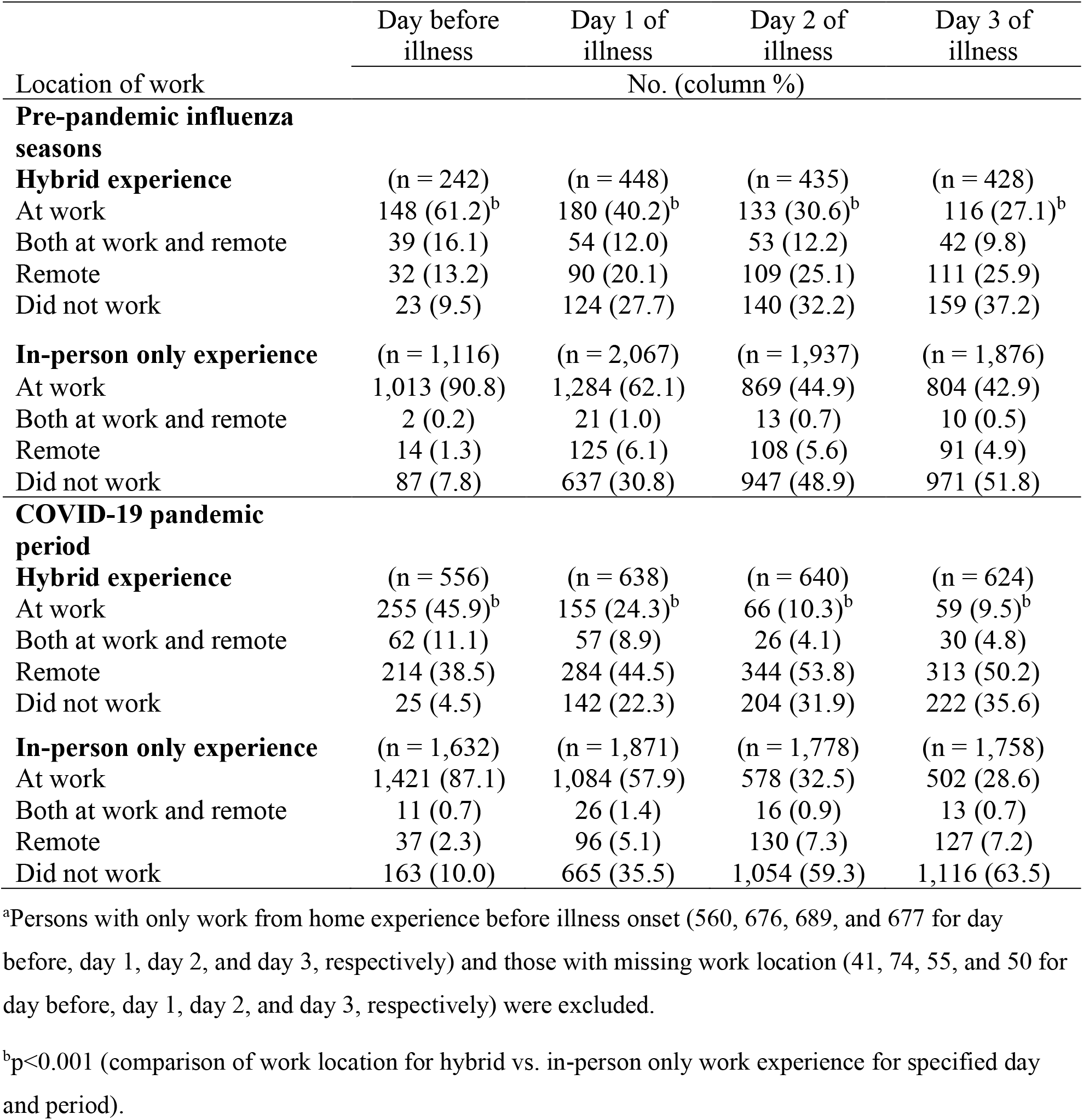
Reported work location among adults with influenza, COVID-19, or other acute respiratory illness who were scheduled to work, by prior experience working from home, United States, 2018-2022^a^.

**Appendix Table 6.**
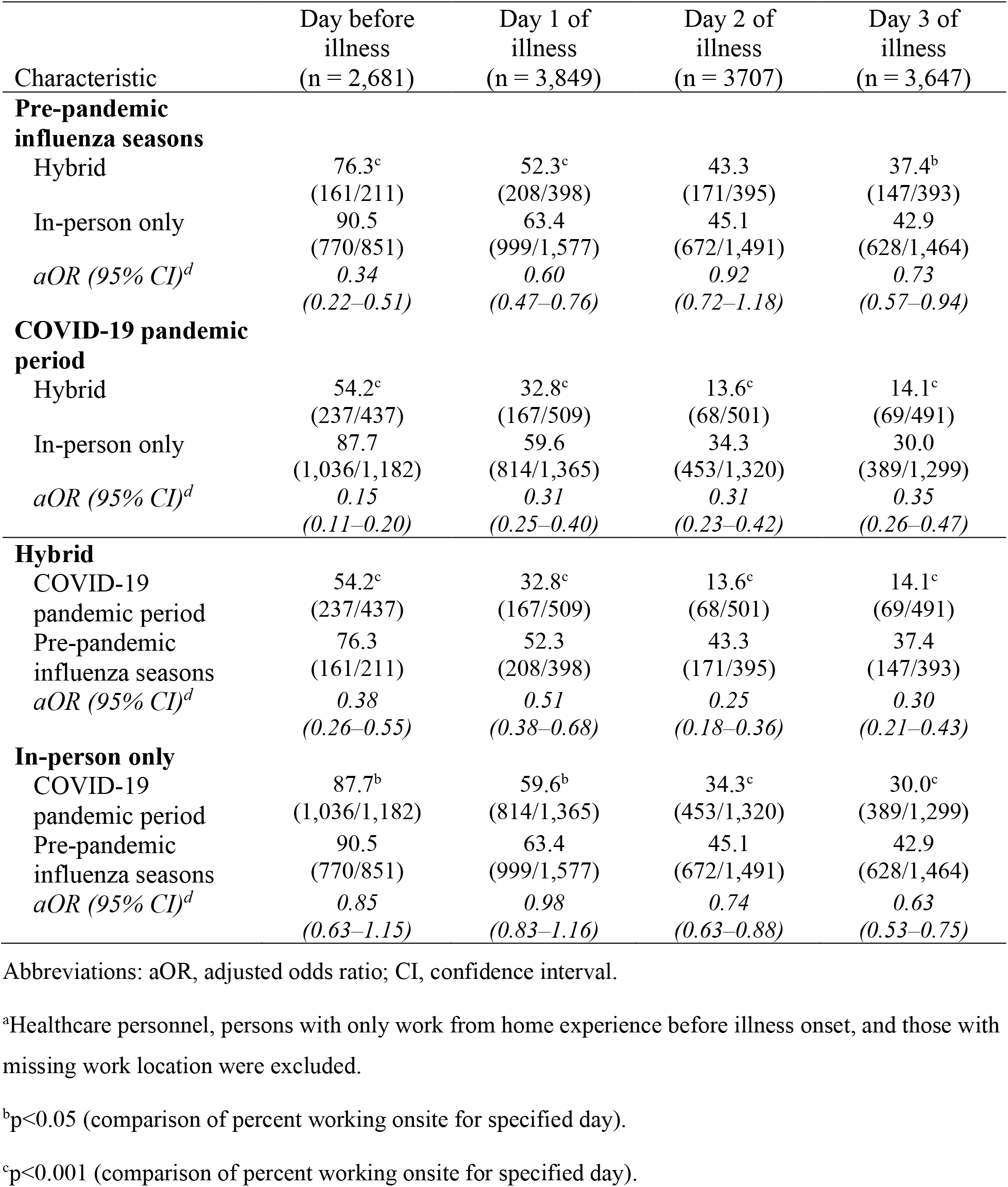
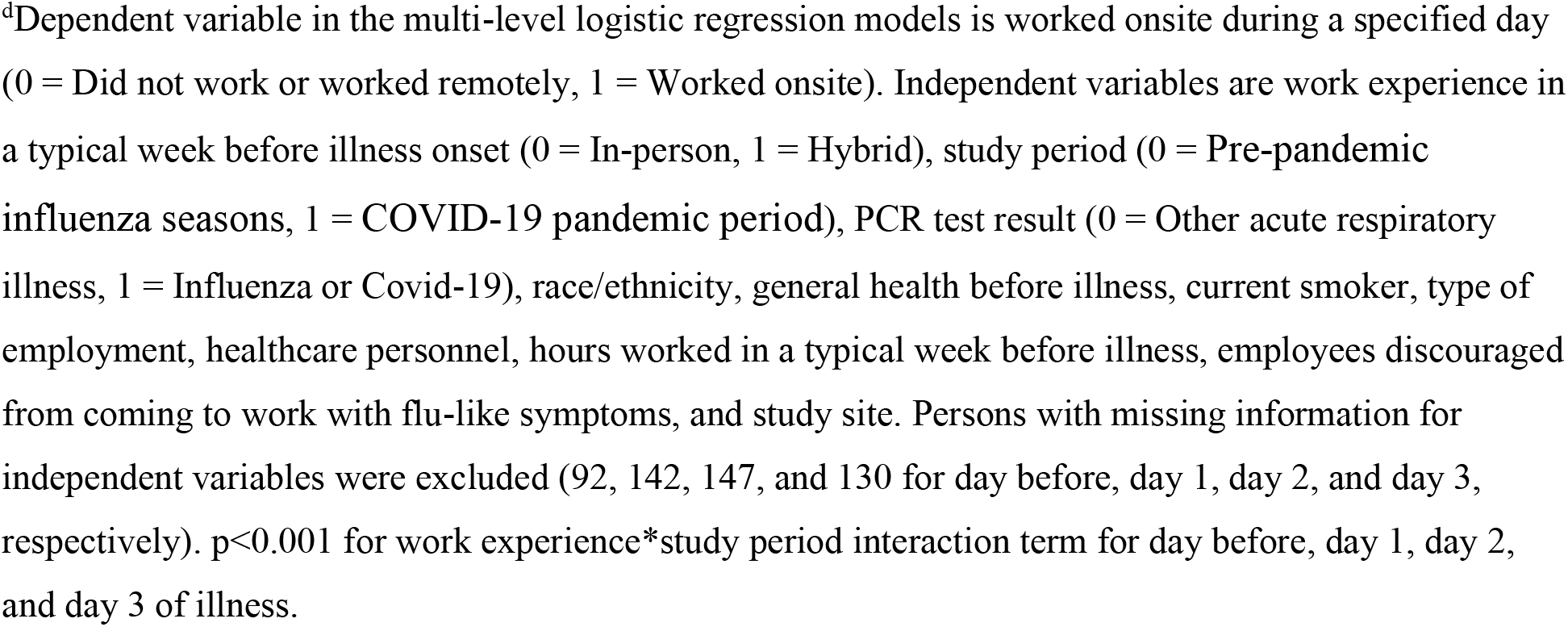
Likelihood of working onsite among *non-healthcare personnel* with COVID-19, influenza, or other acute respiratory illness who were scheduled to work, by prior experience working from home, United States, 2018–2022^a^.

| Characteristic | Day before illness<br>(n = 2,681) | Day 1 of illness<br>(n = 3,849) | Day 2 of illness<br>(n = 3707) | Day 3 of illness<br>(n = 3,647) |
| --- | --- | --- | --- | --- |
| **Pre-pandemic influenza seasons** |  |  |  |  |
| Hybrid | 76.3<sup>c</sup><br>(161/211) | 52.3<sup>c</sup><br>(208/398) | 43.3<br>(171/395) | 37.4<sup>b</sup><br>(147/393) |
| In-person only | 90.5<br>(770/851) | 63.4<br>(999/1,577) | 45.1<br>(672/1,491) | 42.9<br>(628/1,464) |
| *aOR (95% CI)<sup>d</sup>* | *0.34*<br>(0.22–0.51) | *0.60*<br>(0.47–0.76) | *0.92*<br>(0.72–1.18) | *0.73*<br>(0.57–0.94) |
| **COVID-19 pandemic period** |  |  |  |  |
| Hybrid | 54.2<sup>c</sup><br>(237/437) | 32.8<sup>c</sup><br>(167/509) | 13.6<sup>c</sup><br>(68/501) | 14.1<sup>c</sup><br>(69/491) |
| In-person only | 87.7<br>(1,036/1,182) | 59.6<br>(814/1,365) | 34.3<br>(453/1,320) | 30.0<br>(389/1,299) |
| *aOR (95% CI)<sup>d</sup>* | *0.15*<br>(0.11–0.20) | *0.31*<br>(0.25–0.40) | *0.31*<br>(0.23–0.42) | *0.35*<br>(0.26–0.47) |
| **Hybrid** |  |  |  |  |
| COVID-19 pandemic period | 54.2<sup>c</sup><br>(237/437) | 32.8<sup>c</sup><br>(167/509) | 13.6<sup>c</sup><br>(68/501) | 14.1<sup>c</sup><br>(69/491) |
| Pre-pandemic influenza seasons | 76.3<br>(161/211) | 52.3<br>(208/398) | 43.3<br>(171/395) | 37.4<br>(147/393) |
| *aOR (95% CI)<sup>d</sup>* | *0.38*<br>(0.26–0.55) | *0.51*<br>(0.38–0.68) | *0.25*<br>(0.18–0.36) | *0.30*<br>(0.21–0.43) |
| **In-person only** |  |  |  |  |
| COVID-19 pandemic period | 87.7<sup>b</sup><br>(1,036/1,182) | 59.6<sup>b</sup><br>(814/1,365) | 34.3<sup>c</sup><br>(453/1,320) | 30.0<sup>c</sup><br>(389/1,299) |
| Pre-pandemic influenza seasons | 90.5<br>(770/851) | 63.4<br>(999/1,577) | 45.1<br>(672/1,491) | 42.9<br>(628/1,464) |
| *aOR (95% CI)<sup>d</sup>* | *0.85*<br>(0.63–1.15) | *0.98*<br>(0.83–1.16) | *0.74*<br>(0.63–0.88) | *0.63*<br>(0.53–0.75) |

Abbreviations: aOR, adjusted odds ratio; CI, confidence interval.
<sup>a</sup>Healthcare personnel, persons with only work from home experience before illness onset, and those with missing work location were excluded.
<sup>b</sup>p<0.05 (comparison of percent working onsite for specified day).
<sup>c</sup>p<0.001 (comparison of percent working onsite for specified day).
<sup>d</sup>Dependent variable in the multi-level logistic regression models is worked onsite during a specified day (0 = Did not work or worked remotely, 1 = Worked onsite). Independent variables are work experience in a typical week before illness onset (0 = In-person, 1 = Hybrid), study period (0 = Pre-pandemic influenza seasons, 1 = COVID-19 pandemic period), PCR test result (0 = Other acute respiratory illness, 1 = Influenza or Covid-19), race/ethnicity, general health before illness, current smoker, type of employment, healthcare personnel, hours worked in a typical week before illness, employees discouraged from coming to work with flu-like symptoms, and study site. Persons with missing information for independent variables were excluded (92, 142, 147, and 130 for day before, day 1, day 2, and day 3, respectively). $p < 0.001$ for work experience\*study period interaction term for day before, day 1, day 2, and day 3 of illness.

**Appendix Table 7.**
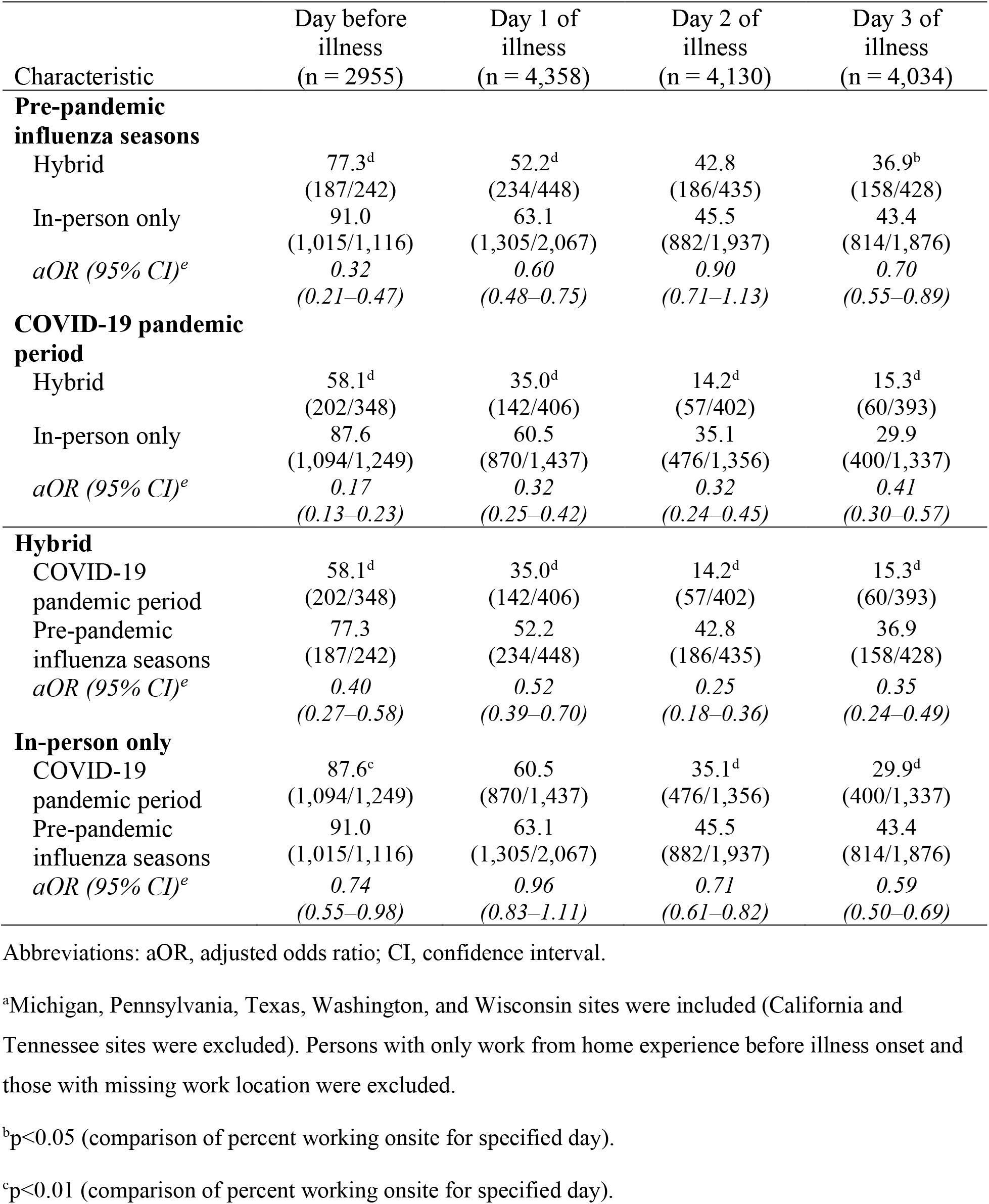
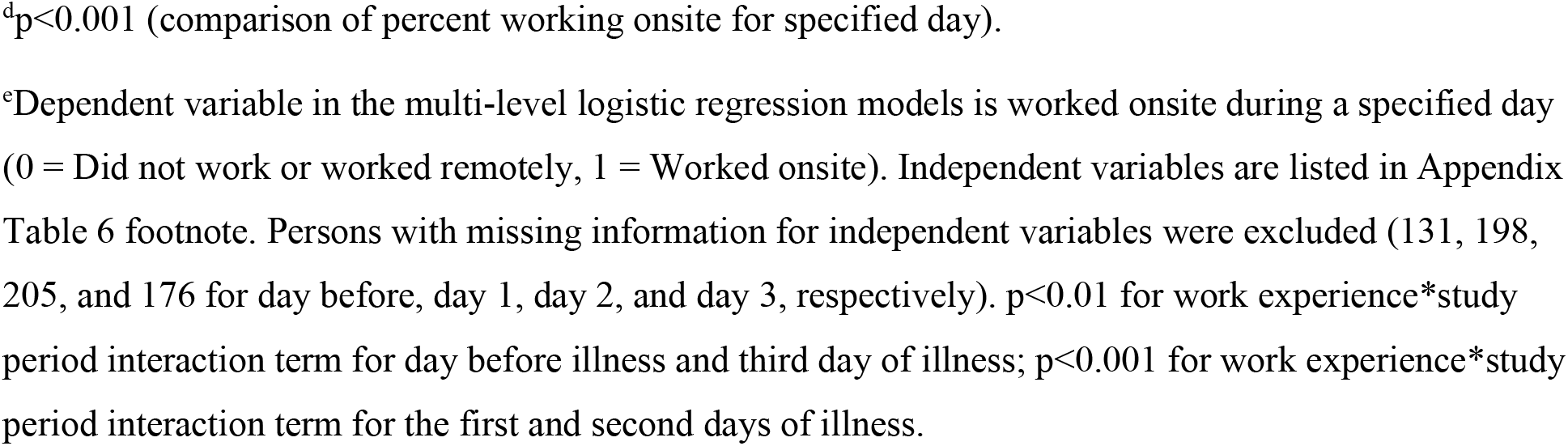
Likelihood of working onsite among adults with COVID-19, influenza, or other acute respiratory illness who were scheduled to work for the *five sites that contributed data for all four years*, by prior experience working from home, United States, 2018–2022^a^.

| Characteristic | Day before illness<br>(n = 2955) | Day 1 of illness<br>(n = 4,358) | Day 2 of illness<br>(n = 4,130) | Day 3 of illness<br>(n = 4,034) |
| --- | --- | --- | --- | --- |
| **Pre-pandemic influenza seasons** |  |  |  |  |
| Hybrid | 77.3<sup>d</sup><br>(187/242) | 52.2<sup>d</sup><br>(234/448) | 42.8<br>(186/435) | 36.9<sup>b</sup><br>(158/428) |
| In-person only | 91.0<br>(1,015/1,116) | 63.1<br>(1,305/2,067) | 45.5<br>(882/1,937) | 43.4<br>(814/1,876) |
| *aOR (95% CI)<sup>e</sup>* | *0.32*<br>*(0.21–0.47)* | *0.60*<br>*(0.48–0.75)* | *0.90*<br>*(0.71–1.13)* | *0.70*<br>*(0.55–0.89)* |
| **COVID-19 pandemic period** |  |  |  |  |
| Hybrid | 58.1<sup>d</sup><br>(202/348) | 35.0<sup>d</sup><br>(142/406) | 14.2<sup>d</sup><br>(57/402) | 15.3<sup>d</sup><br>(60/393) |
| In-person only | 87.6<br>(1,094/1,249) | 60.5<br>(870/1,437) | 35.1<br>(476/1,356) | 29.9<br>(400/1,337) |
| *aOR (95% CI)<sup>e</sup>* | *0.17*<br>*(0.13–0.23)* | *0.32*<br>*(0.25–0.42)* | *0.32*<br>*(0.24–0.45)* | *0.41*<br>*(0.30–0.57)* |
| **Hybrid** |  |  |  |  |
| COVID-19 pandemic period | 58.1<sup>d</sup><br>(202/348) | 35.0<sup>d</sup><br>(142/406) | 14.2<sup>d</sup><br>(57/402) | 15.3<sup>d</sup><br>(60/393) |
| Pre-pandemic influenza seasons | 77.3<br>(187/242) | 52.2<br>(234/448) | 42.8<br>(186/435) | 36.9<br>(158/428) |
| *aOR (95% CI)<sup>e</sup>* | *0.40*<br>*(0.27–0.58)* | *0.52*<br>*(0.39–0.70)* | *0.25*<br>*(0.18–0.36)* | *0.35*<br>*(0.24–0.49)* |
| **In-person only** |  |  |  |  |
| COVID-19 pandemic period | 87.6<sup>c</sup><br>(1,094/1,249) | 60.5<br>(870/1,437) | 35.1<sup>d</sup><br>(476/1,356) | 29.9<sup>d</sup><br>(400/1,337) |
| Pre-pandemic influenza seasons | 91.0<br>(1,015/1,116) | 63.1<br>(1,305/2,067) | 45.5<br>(882/1,937) | 43.4<br>(814/1,876) |
| *aOR (95% CI)<sup>e</sup>* | *0.74*<br>*(0.55–0.98)* | *0.96*<br>*(0.83–1.11)* | *0.71*<br>*(0.61–0.82)* | *0.59*<br>*(0.50–0.69)* |

Abbreviations: aOR, adjusted odds ratio; CI, confidence interval.
<sup>a</sup>Michigan, Pennsylvania, Texas, Washington, and Wisconsin sites were included (California and Tennessee sites were excluded). Persons with only work from home experience before illness onset and those with missing work location were excluded.
<sup>b</sup>p<0.05 (comparison of percent working onsite for specified day).
<sup>c</sup>p<0.01 (comparison of percent working onsite for specified day).
<sup>d</sup>p<0.001 (comparison of percent working onsite for specified day).
<sup>e</sup>Dependent variable in the multi-level logistic regression models is worked onsite during a specified day (0 = Did not work or worked remotely, 1 = Worked onsite). Independent variables are listed in Appendix Table 6 footnote. Persons with missing information for independent variables were excluded (131, 198, 205, and 176 for day before, day 1, day 2, and day 3, respectively). p<0.01 for work experience\*study period interaction term for day before illness and third day of illness; p<0.001 for work experience\*study period interaction term for the first and second days of illness.

**Appendix Table 8.**
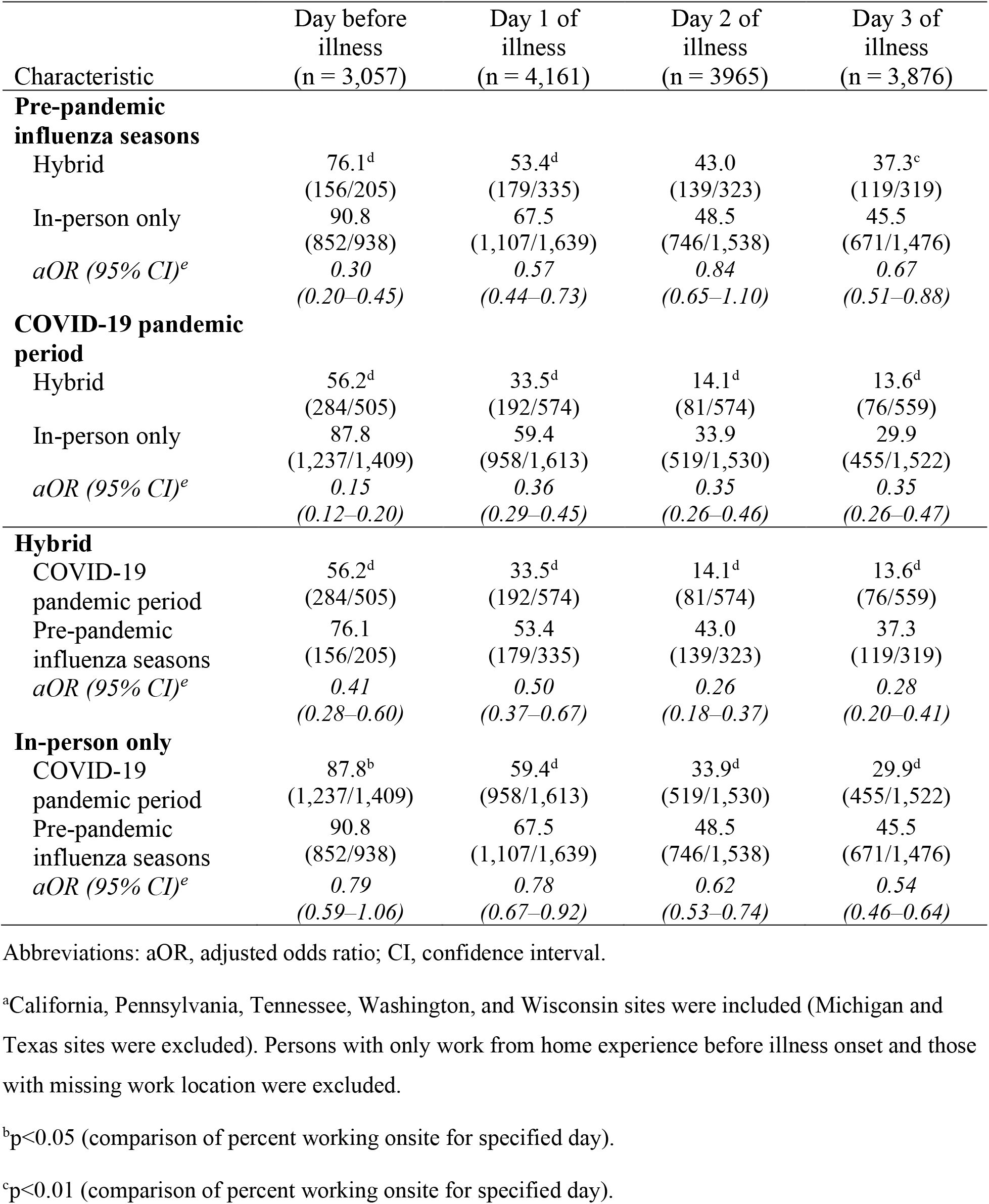
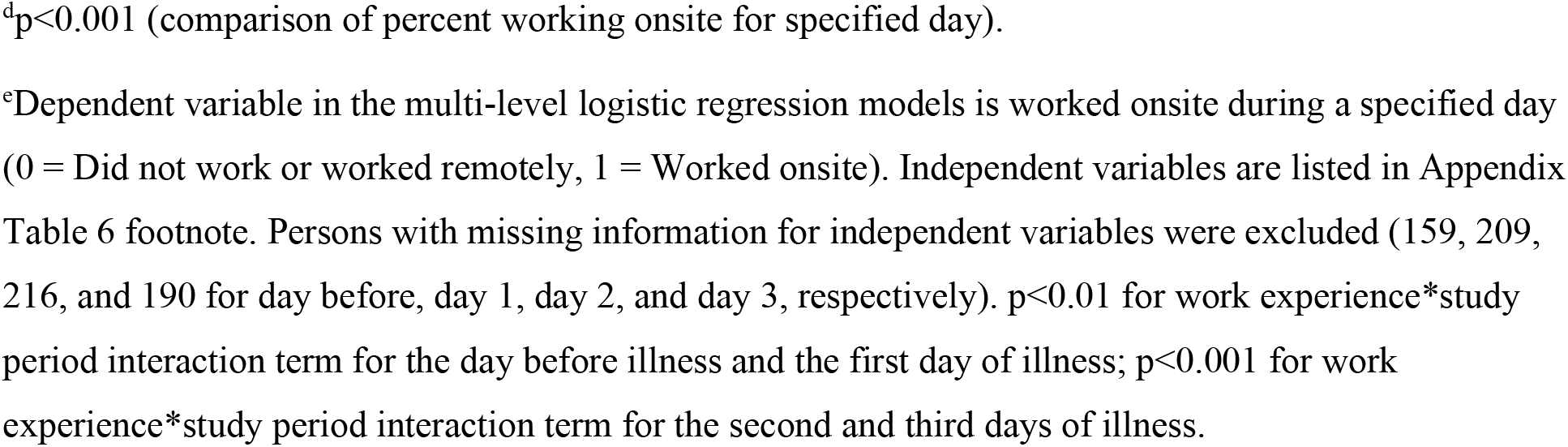
Likelihood of working onsite among adults with COVID-19, influenza, or other acute respiratory illness who were scheduled to work for the *five sites with higher survey completion rates*, by prior experience working from home, United States, 2018–2022^a^.

| Characteristic | Day before illness<br>(n = 3,057) | Day 1 of illness<br>(n = 4,161) | Day 2 of illness<br>(n = 3965) | Day 3 of illness<br>(n = 3,876) |
| --- | --- | --- | --- | --- |
| **Pre-pandemic influenza seasons** |  |  |  |  |
| Hybrid | 76.1<sup>d</sup><br>(156/205) | 53.4<sup>d</sup><br>(179/335) | 43.0<br>(139/323) | 37.3<sup>c</sup><br>(119/319) |
| In-person only | 90.8<br>(852/938) | 67.5<br>(1,107/1,639) | 48.5<br>(746/1,538) | 45.5<br>(671/1,476) |
| *aOR (95% CI)<sup>e</sup>* | *0.30*<br>(0.20–0.45) | *0.57*<br>(0.44–0.73) | *0.84*<br>(0.65–1.10) | *0.67*<br>(0.51–0.88) |
| **COVID-19 pandemic period** |  |  |  |  |
| Hybrid | 56.2<sup>d</sup><br>(284/505) | 33.5<sup>d</sup><br>(192/574) | 14.1<sup>d</sup><br>(81/574) | 13.6<sup>d</sup><br>(76/559) |
| In-person only | 87.8<br>(1,237/1,409) | 59.4<br>(958/1,613) | 33.9<br>(519/1,530) | 29.9<br>(455/1,522) |
| *aOR (95% CI)<sup>e</sup>* | *0.15*<br>(0.12–0.20) | *0.36*<br>(0.29–0.45) | *0.35*<br>(0.26–0.46) | *0.35*<br>(0.26–0.47) |
| **Hybrid** |  |  |  |  |
| COVID-19 pandemic period | 56.2<sup>d</sup><br>(284/505) | 33.5<sup>d</sup><br>(192/574) | 14.1<sup>d</sup><br>(81/574) | 13.6<sup>d</sup><br>(76/559) |
| Pre-pandemic influenza seasons | 76.1<br>(156/205) | 53.4<br>(179/335) | 43.0<br>(139/323) | 37.3<br>(119/319) |
| *aOR (95% CI)<sup>e</sup>* | *0.41*<br>(0.28–0.60) | *0.50*<br>(0.37–0.67) | *0.26*<br>(0.18–0.37) | *0.28*<br>(0.20–0.41) |
| **In-person only** |  |  |  |  |
| COVID-19 pandemic period | 87.8<sup>b</sup><br>(1,237/1,409) | 59.4<sup>d</sup><br>(958/1,613) | 33.9<sup>d</sup><br>(519/1,530) | 29.9<sup>d</sup><br>(455/1,522) |
| Pre-pandemic influenza seasons | 90.8<br>(852/938) | 67.5<br>(1,107/1,639) | 48.5<br>(746/1,538) | 45.5<br>(671/1,476) |
| *aOR (95% CI)<sup>e</sup>* | *0.79*<br>(0.59–1.06) | *0.78*<br>(0.67–0.92) | *0.62*<br>(0.53–0.74) | *0.54*<br>(0.46–0.64) |

Abbreviations: aOR, adjusted odds ratio; CI, confidence interval.
<sup>a</sup>California, Pennsylvania, Tennessee, Washington, and Wisconsin sites were included (Michigan and Texas sites were excluded). Persons with only work from home experience before illness onset and those with missing work location were excluded.
<sup>b</sup>p<0.05 (comparison of percent working onsite for specified day).
<sup>c</sup>p<0.01 (comparison of percent working onsite for specified day).
<sup>d</sup> $p < 0.001$ (comparison of percent working onsite for specified day).
<sup>e</sup>Dependent variable in the multi-level logistic regression models is worked onsite during a specified day (0 = Did not work or worked remotely, 1 = Worked onsite). Independent variables are listed in Appendix Table 6 footnote. Persons with missing information for independent variables were excluded (159, 209, 216, and 190 for day before, day 1, day 2, and day 3, respectively). $p < 0.01$ for work experience\*study period interaction term for the day before illness and the first day of illness; $p < 0.001$ for work experience\*study period interaction term for the second and third days of illness.

**Appendix Table 9.** Reported work location among adults with influenza, COVID-19, or other acute respiratory illness (ARI) who were scheduled to work, by PCR test result, United States, 2018-2022^a^.

|  | Day before<br>illness | Day 1 of<br>illness | Day 2 of<br>illness | Day 3 of<br>illness |
| --- | --- | --- | --- | --- |
| Location of work | No. (column percent) |  |  |  |
| **Pre-pandemic influenza seasons** |  |  |  |  |
| **Influenza** | (n = 499) | (n = 850) | (n = 835) | (n = 837) |
| At work | 430 (86.2) | 481 (56.6)<sup>b</sup> | 272 (32.6)<sup>c</sup> | 227 (27.1)<sup>c</sup> |
| Both at work and remote | 13 (2.6) | 23 (2.7) | 13 (1.5) | 9 (1.1) |
| Remote | 13 (2.6) | 59 (6.9) | 69 (8.3) | 62 (7.4) |
| Did not work | 43 (8.6) | 287 (33.8) | 481 (57.6) | 539 (64.4) |
| **Other ARI** |  |  |  |  |
|  | (n = 859) | (n = 1,665) | (n = 1,537) | (n = 1,467) |
| At work | 731 (85.1) | 983 (59.0) | 730 (47.5) | 693 (47.3) |
| Both at work and remote | 28 (3.3) | 52 (3.1) | 53 (3.5) | 43 (2.9) |
| Remote | 33 (3.8) | 156 (9.4) | 148 (9.6) | 140 (9.5) |
| Did not work | 67 (7.8) | 474 (28.5) | 606 (39.4) | 591 (40.3) |
| **COVID-19 pandemic period** |  |  |  |  |
| **COVID-19** | (n = 865) | (n = 1,020) | (n = 986) | (n = 974) |
| At work | 656 (75.8)<sup>b</sup> | 488 (47.9) | 209 (21.2)<sup>c</sup> | 124 (12.8)<sup>c</sup> |
| Both at work and remote | 25 (2.9) | 34 (3.3) | 11 (1.1) | 13 (1.3) |
| Remote | 94 (10.9) | 161 (15.8) | 197 (20.0) | 187 (19.2) |
| Did not work | 90 (10.4) | 337 (33.0) | 569 (57.7) | 650 (66.7) |
| **Other ARI** |  |  |  |  |
|  | (n = 1,266) | (n = 1,424) | (n = 1,362) | (n = 1,341) |
| At work | 984 (77.7) | 719 (50.5) | 419 (30.7) | 422 (31.5) |
| Both at work and remote | 47 (3.7) | 49 (3.4) | 31 (2.3) | 30 (2.2) |
| Remote | 148 (11.7) | 209 (14.7) | 265 (19.5) | 241 (18.0) |
| Did not work | 87 (6.9) | 447 (31.4) | 647 (47.5) | 648 (48.3) |

<sup>a</sup>Persons with influenza during Oct 2020–Jun 2022 (57, 65, 70, and 67 for day before, day 1, day 2, and day 3, respectively), persons with only work from home experience before illness onset (560, 676, 689, and 677 for day before, day 1, day 2, and day 3, respectively), and persons with missing work location (41, 74, 55, and 50 for day before, day 1, day 2, and day 3, respectively) were excluded.
<sup>b</sup>p<0.05 (comparison of work location by test result for specified day and period).
<sup>c</sup>p<0.001 (comparison of work location by test result for specified day and period).

**Appendix Table 10.** Likelihood of working onsite among adults with COVID-19 illness who were scheduled to work, by day when COVID-19 positive PCR and at-home test results were available, United States, *January 2022–June 2022*^a^.

| Characteristic | Worked onsite,<br>% |
| --- | --- |
| Scheduled to work on day 1 of COVID-19 illness<sup>b</sup> |  |
| COVID-19 positive PCR result available on day 1 of illness | 83.3 (5/6) |
| COVID-19 positive PCR result available after day 1 of illness | 50.3 (173/344) |
| COVID-19 positive at-home result available on day 1 of illness | 38.7 (12/31) |
| Excluding persons in the category above | 51.4 (161/313) |
| Scheduled to work on day 2 of COVID-19 illness<sup>c</sup> |  |
| COVID-19 positive PCR result available on day 1 or 2 of illness | 0.0 (0/56)<sup>f</sup> |
| COVID-19 positive PCR result available after day 2 of illness | 18.6 (52/280) |
| COVID-19 positive at-home result available on day 1 or 2 of illness | 11.1 (7/63) |
| Excluding persons in the category above | 20.7 (45/217) |
| Scheduled to work on day 3 of COVID-19 illness<sup>d</sup> |  |
| COVID-19 positive PCR result available on day 1, 2, or 3 of illness | 3.5 (4/116)<sup>e</sup> |
| COVID-19 positive PCR result available after day 3 of illness | 12.0 (26/216) |
| COVID-19 positive at-home result available on day 1, 2, or 3 of illness | 11.3 (7/62) |
| Excluding persons in the category above | 12.3 (19/154) |

Values represent percent worked onsite (at work + both at work and remotely / scheduled to work).
Analysis is based on persons with COVID-19 shown in Table 5 who enrolled in the study on or after January 15, 2022.
<sup>a</sup>Day of illness when COVID-19 positive result was available was computed by comparing the date of illness onset with the date that COVID-19 positive test result was available.
<sup>b</sup>Unknown when COVID-19 positive PCR result was available = 16 persons.
<sup>c</sup>Unknown when COVID-19 positive PCR result was available = 19 persons.
<sup>d</sup>Unknown when COVID-19 positive PCR result was available = 18 persons.
<sup>e</sup>p<0.01 (comparison of the two PCR result categories for specified day of illness).
<sup>f</sup>p<0.001 (comparison of the two PCR result categories for specified day of illness).

**Appendix Figure 1.**
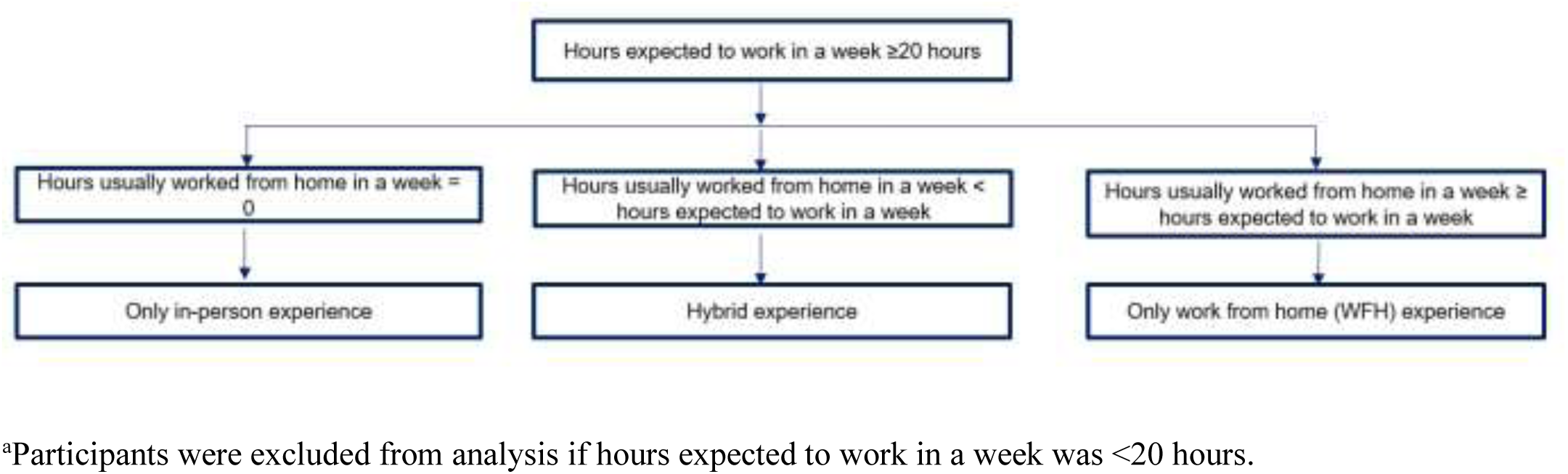
Algorithm to categorize work experience before illness onset, United States, 2018–2022^a^.

**Appendix Figure 2.**
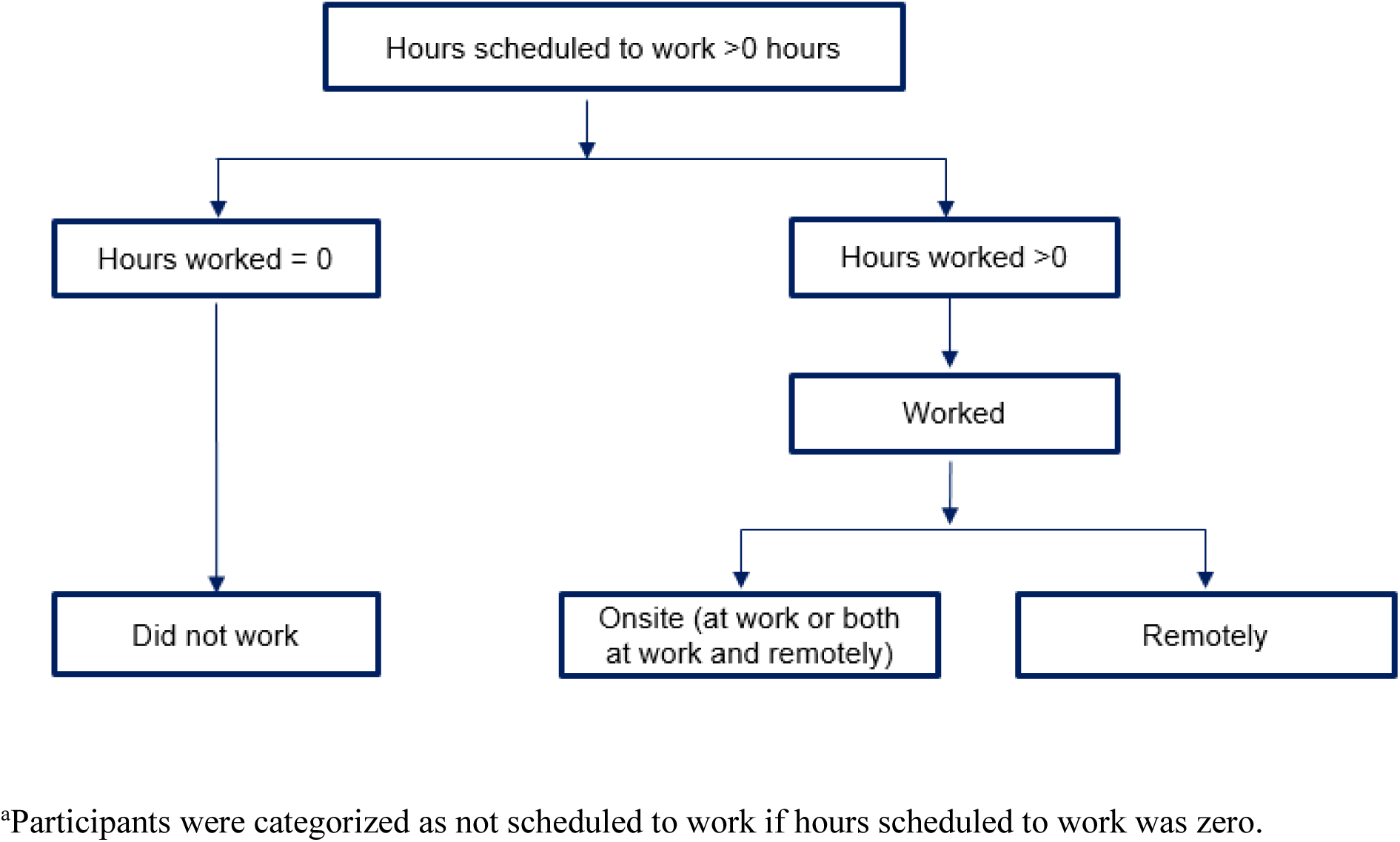
Algorithm to categorize work attendance on the day before illness onset and during the first 3 days of illness among persons who were scheduled to work, United States, 2018–2022^a^.

**Appendix Figure 3.**
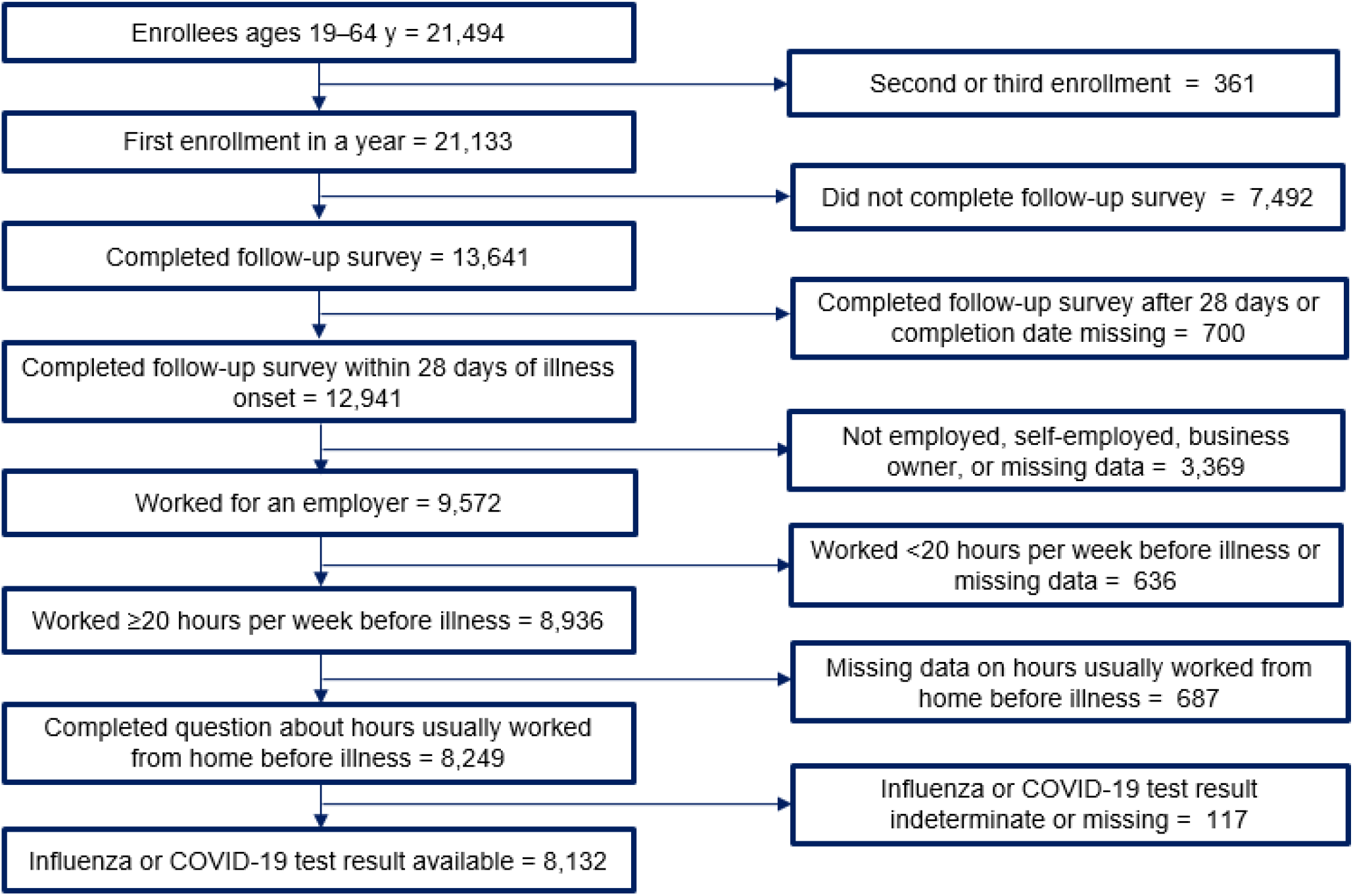
Assembly of study participants with influenza, COVID-19, or other acute respiratory illness, United States, 2018–2022.

**Appendix Figure 4.**
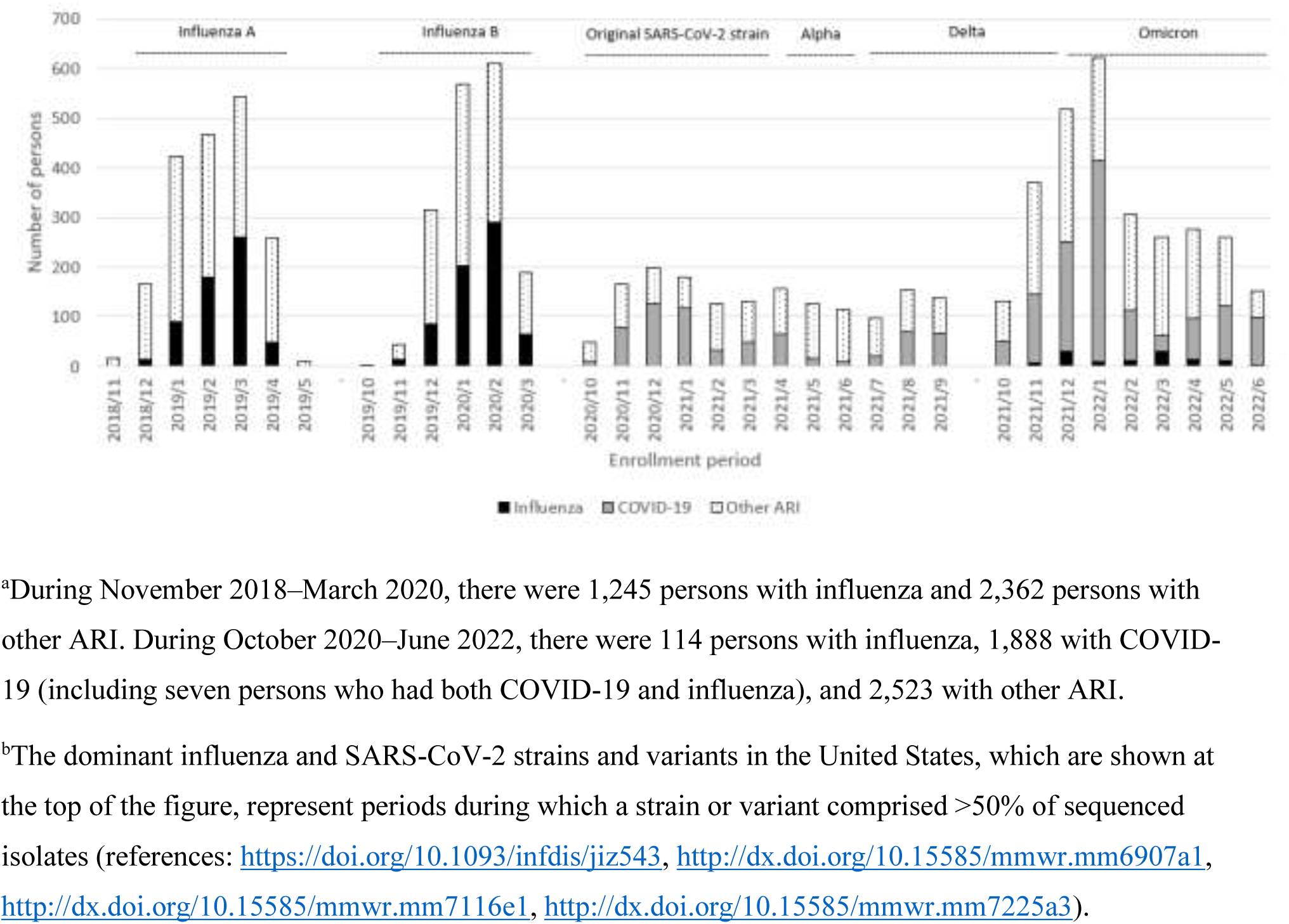
Period of enrollment of adults with influenza, COVID-19, or other acute respiratory illness (ARI) who were included in the analysis (n = 8,132), United States, 2018– 2022^a,b^.

## Acknowledgements

This work was supported through cooperative agreements funded by the US Centers for Disease Control and Prevention and by infrastructure funding from the National Institutes of Health (UL1 TR001857) at the University of Pittsburgh. We gratefully acknowledge the contributions of the following persons: Chandni Raiyani, Kayan Dunnigan, Kempapura Murthy, Mufaddal Mamawala, Spencer Rose, Amanda McKillop, Teresa Ponder, Ashley Graves, Martha Zayed, Natalie Settele, Jason Ettlinger, Courtney Shaver, Monica Bennett, Elisa Priest, Jennifer Thomas, Eric Hoffman, Marcus Volz, Kimberly Walker, Manohar Mutnal, Arundhati Rao, Michael Reis, Keith Stone, Madhava Beeram, and Alejandro Arroliga. Krissy Moehling Geffel, Rachel Taber, Jonathan Raviotta, Louise Taylor, Michael Susick, GK Balasubramani, Theresa M. Sax. Vanderbilt University Medical Center: Dayna Wyatt, Stephanie Longmire, Meredith Denny, Zhouwen Liu, Yuwei Zhu. Sally Shaw, Jeniffer Kim. Edward Belongia, Hannah Berger, Jennifer Meece, Carla Rottscheit, Erik Kronholm, Jackie Salzwedel, Julie Karl, Anna Zachow, Linda Heeren, Joshua Blake, Jennifer Moran, Christopher Rayburn, Stephanie Kohl, Christian Delgadillo, Vicki Moon, Megan Tichenor, Miriah Rotar, Kelly Scheffen. Erika Kiniry, Stacie Wellwood, Brianna Wickersham, Matt Nguyen, Rachael Doud, Suzie Park, Mike Jackson.

## Disclaimer

The findings and conclusions are those of the authors and do not necessarily represent the official position of the Centers for Disease Control and Prevention.

## About the Author

Dr. Ahmed is a senior science officer at the Division of Global Migration and Quarantine, National Center for Emerging and Zoonotic Infectious Diseases, Centers for Disease Control and Prevention, Atlanta, Georgia, USA. His research interests include prevention and control of emerging infectious diseases, including pandemic influenza and COVID-19.

